# Brain Metabolic Activity on FDG PET/CT Predicts Survival in Non–Small Cell Lung Cancer

**DOI:** 10.1101/2025.10.07.25337046

**Authors:** Julie Auriac, Ghada Lemoudda, Narinée Hovhannisyan-Baghdasarian, Manuel Pires, Lalith Kumar Shiyam Sundar, Paulette Salamoun-Feghali, Romain-David Seban, Nina Jehanno, Christophe Nioche, Marie Luporsi, Thomas Beyer, Alain Livartowski, Nicolas Girard, Irène Buvat, Fanny Orlhac

## Abstract

**Background:** [18F]FDG-PET/CT images play a key role in the management of patients with Non-Small Cell Lung Cancer (NSCLC). However, in these scans, the focus is always on the detected tumors and on their characteristics, neglecting information from other organs or tissues.

**Purpose:** To investigate whether the mean brain FDG uptake (SUVmean_brain_) is associated with overall survival (OS) in metastatic NSCLC patients.

**Materials and Methods:** This retrospective study included metastatic NSCLC patients who underwent pre-treatment [18F]FDG-PET/CT scans between 2010 and 2023. Clinical and biological data, tumor radiomic features, and SUVmean_brain_ were collected. The ability of these features to predict OS was evaluated using univariable and multivariable Cox regression models. The correlation between SUVmean_brain_ and clinical/imaging/blood biomarkers was investigated using Spearman correlation coefficients (rS).

**Results:** Patients were chronologically divided into a discovery (n = 234, mean age 64 years ± 11 [SD], 135 male) and test set (n = 146, mean age 66 years ± 11, 83 male). In the discovery set, univariable analysis showed that high SUVmean_brain_ (≥ median) was associated with longer OS (HR = 0.83, 95% CI 0.76-0.92, *P* < .001). SUVmean_brain_ was significantly lower in patients who died within one year compared to those who were still alive (*P* < .001). In multivariable analysis, SUVmean_brain_ remained an independent prognostic factor for OS (HR, 0.88; 95% CI: 0.80, 0.98; *P* = .02), which was confirmed in the test set (*P* < .001). SUVmean_brain_ was independent of the radiomic features quantifying tumor involvement (|rS| < 0.24, n = 380 patients) and significantly correlated but complementary to several blood biomarkers including CRP (rS = -0.37, n = 110 patients).

**Conclusion:** Low brain metabolic activity was associated with increased mortality in metastatic NSCLC patients. SUVmean_brain_ was an independent prognostic factor that could contribute to patient stratification, although its interpretation requires further investigation.

## Introduction

Mortality from lung cancer is the highest among cancers with a total of 1.8 million deaths (18%) worldwide in 2022^1^. Despite promising new therapies, the prognosis for patients is relatively poor with overall survival ranging from 37.5 months in the early stages to 4.8 months in the metastatic stage^2^. In clinical practice, the management of metastatic lung cancer patients remains a challenge, as some patients are resistant to treatments, or only experience short-term benefits, without any significant impact on their survival^3–4^.

18F-fluorodeoxyglucose ([18F]FDG) positron emission tomography/computed tomography (PET/CT) imaging of lung cancer patients has proven valuable for the diagnosis, staging and evaluation of treatment^5^. Numerous studies have shown that advanced analysis of whole-body [18F]FDG PET/CT images could provide valuable information from the tumor invasion to predict patient survival in lung cancer. Total Metabolic Tumor Volume (TMTV), Total Lesion Glycolysis (TLG), maximum Standardized Uptake Value (SUVmax) or the maximum distance between two lesions (Dmax) are PET-derived biomarkers reflecting the metabolic tumor burden, tumor activity or dissemination, with a strong prognostic value for patient survival^6–10^.

Recent studies suggest that metabolic activity and radiodensity in various organs and tissues, measured from whole-body [18F]FDG PET/CT images, provide additional valuable information for predicting patients’ outcomes in lung cancer^11–14^, by reflecting the patient’s general condition. FDG uptake measured in lymphoid organs may reflect systemic inflammation, and has been associated with prognosis in advanced NSCLC^15–17^. Systemic inflammation, often characterized by blood-based biomarkers such as C-reactive protein (CRP), has been associated with treatment response and prognosis in lung cancer^18–20^. More recently, nutritional and immune status have been introduced sas potential prognostic factors in cancer progression and outcomes in patients with lung cancer^21–22^.

In parallel, several publications suggest the existence of a lung-brain axis^23^, indicating that diseases affecting the pulmonary system can lead to alterations in brain structure and function. In particular, brain alterations have been observed in patients with lung cancer compared to healthy controls, including prior to the initiation of chemotherapy^24–27^. In addition, recent studies have suggested a link between neuroinflammation and the pathogenesis and cachexia^28^, between tumor growth and neuroinflammation^29^, and between neuroinflammation and systemic inflammation^30^, calling for further investigations regarding the role of the brain function in cancer patients.

In this context, the purpose of our study was to investigate a potential relationship between brain metabolic activity as measured from [18F]FDG PET and survival in metastatic non-small cell lung cancer.

## Materials and Methods

This study was approved by the institutional review board of Institut Curie (DATA220207). Informed consent was obtained for all patients through institutional processes. All related data were de-identified, collected and stored in compliance with GDPR and the Declaration of Helsinki.

### 1. Patients

This retrospective study included 455 patients with metastatic NSCLC treated at Institut Curie between 2010 and 2023, who performed baseline [18F]FDG PET/CT before any treatment. Patients were treated according to the standard of care at the time of their management. The exclusion criteria were as follows: a) patients with no baseline [18F]FDG PET/CT imaging (i.e. surgery or neoadjuvant therapy prior imaging), b) follow-up duration lower than 12 months, c) missing clinical data, d) brain not fully included in the axial [18F]FDG PET/CT field of view, and e) no lesion with an SUV greater than 4. Clinical data composed of age, sex, Body Mass Index (BMI), smoking history, performance status (PS), and treatment administered were collected. Blood parameters including albumin, C-reactive protein (CRP), monocytes, leukocytes, lymphocytes, neutrophils and cortisol were collected when available within 30 days of the PET scan and prior treatment initiation. The Prognostic Nutritional Index (PNI) defined as albumin (g/L) + 5 × lymphocyte count (10^9^/L) was calculated. Glycaemic control was performed immediately before the PET scan. Patients’ history of depression was also collected retrospectively from the patient medical records when the information was available.

The cohort was divided into two datasets: patients who performed their PET/CT scan between January 2010 and December 2018 were included in the “discovery” set, whereas the others (January 2019 and December 2023) were included in the test set (Figure 1).

**Figure 1:**
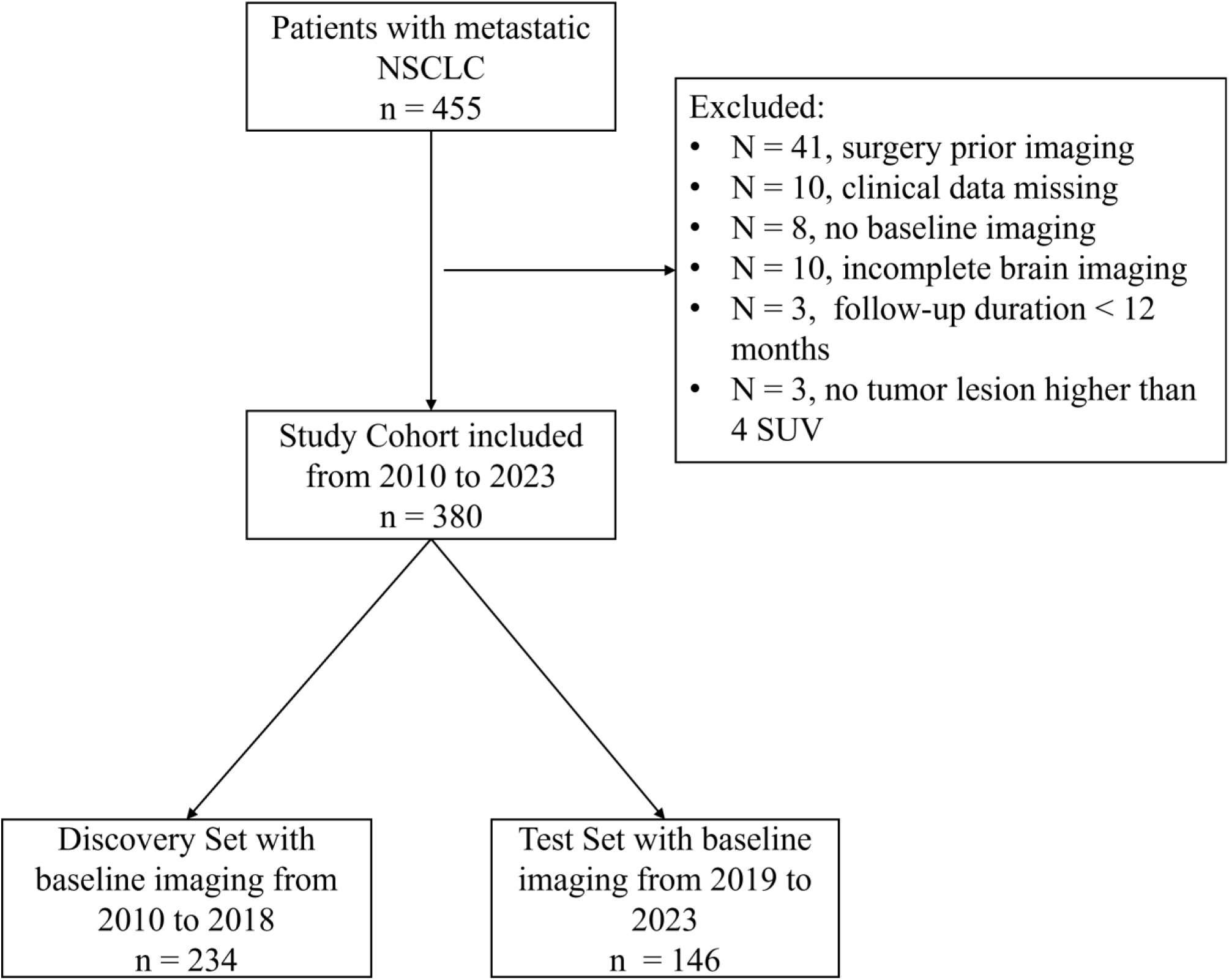
Flow diagram of metastatic NSCLC patients included in our study. The eligible cohort (n = 380) was split chronologically according to the date of PET/CT imaging into a discovery and a test set. The discovery set (n = 234 [62%]) included patients scanned between 2010 and 2018 while the test set (n = 146 [38%]) included patients scanned between 2019 and 2023.

Overall survival was defined as the time between the acquisition of the baseline PET/CT scan and the date of the patient’s death or the last time the patient was known to be alive. Follow-up was determined from the date of the baseline PET/CT scan to the date of the last clinical consultation.

### 2. Imaging and image analysis

#### 2.1 PET/CT imaging

[18F]FDG PET/CT scans were performed at various centres using different PET/CT systems and acquisition and reconstruction protocols (Table S1). The average blood glucose levels before [18F]FDG PET/CT acquisition was (5.96 ± 1.43) mmol/L. An injected dose of (235 ± 75) MBq of [18F]FDG was administered (66 ± 11) minutes before the image acquisition with (1.8 ± 0.7) min per bed position. PET images were expressed in standardized uptake value (SUV) units normalized by the patient body weight.

#### 2.2 Features extraction

For each patient, all tumor lesions were automatically segmented on PET images using LION v.0.14^31^. The lesion delineation was then automatically refined by including only voxels with an SUV equal to or greater than 4 in the tumor region. The whole brain was automatically delineated on the CT images with TotalSegmentator v.2.0.5 software^32^ (https://github.com/wasserth/TotalSegmentator) and the resulting volume of interest (VOI) was transposed to the corresponding PET image (VOI_brain).

Brain metastases identified by any imaging modality (e.g., CT, MRI) at the time of the PET/CT scan or within three months thereafter were manually delineated on the PET images, using the anatomical modality if necessary, and excluded from VOI_brain by a nuclear medicine physician, resulting in a volume corresponding to the whole brain with metastatic lesions removed (VOI_brain_noM).

1. 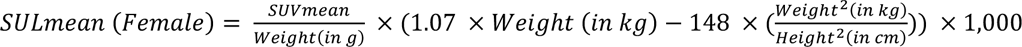
2. 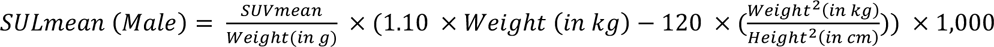

### 3. Statistical analysis

#### Patient Characteristics

Baseline patient characteristics were analysed using the Wilcoxon test for continuous variables and the χ^2^ tests for categorical variables. Spearman’s correlation coefficients (rS) were calculated to assess the correlation between imaging features.

#### Evaluation of automatic segmentation

To evaluate the reliability of the automatic segmentation performed by LION, we compared it to the segmentation performed manually with a SUV 4 threshold by an expert physicist (N.H-B., 10 years of experience) under the supervision of a nuclear physician (M.L., more than 10 years of experience), for 100 randomly selected patients. The DICE score was calculated between the expert and automated segmentations and a Bland-Altman analysis was performed to compare the five feature values as a function of the delineation method.

#### Survival Analysis

Associations between feature values and overall survival (OS) in the discovery set were studied using univariable Cox regression. All variables associated with OS at the *P* < .05 level in univariable analysis were included in a Cox multivariable model, without and with the brain FDG uptake (Model 1 and Model 2 respectively). An analysis of variance (ANOVA) was conducted to compare the two models. A risk score was generated from the multivariable models and then binarized by the median on the discovery set to distinguish high from low risk-patients according to OS. Finally, we applied the same models (same coefficients, same cut-off) to the test set. For both discovery and test sets, the association of low-risk and high-risk groups with OS was evaluated using Kaplan Meier curves with the log-rank test and using a Cox proportional hazard regression model, including calculation of adjusted hazard ratios (HRs) with 95% confidence intervals (CI) and Wald test *P*-values.

#### Identification of factors influencing cerebral metabolic activity

The influence of the presence of brain metastases on the prognostic value of cerebral metabolic activity was investigated by comparing the results obtained with VOI_brain and VOI_brain_noM. The prognostic value of cerebral metabolic activity was also assessed in the subgroup of patients without known brain metastases. The impact of the treatment on the prognostic value of cerebral metabolic activity was studied by separating the patients according to whether they received chemotherapy, radiotherapy, immunotherapy and/or targeted therapy after PET/CT. We used the Wilcoxon test and Spearman correlation coefficient to study the relationship between cerebral metabolic activity and the presence of cerebral metastases, the technical PET/CT acquisition parameters, the value of biological parameters and patients’ depression history. We explored whether integrating cerebral metabolic activity with blood biomarkers could enhance prognostic stratification. For each biological parameter identified as prognostic, patients were stratified into three risk groups according to OS based on the presence of risk factors, defined as cerebral metabolic activity and biological values: low risk (0 risk factor), intermediate risk (1 risk factor) and high risk (2 risk factors). A p-value lower than .05 was defined as statistically significant. All statistical analyses were conducted using R software (R Core Team 2021) version 4.2.2.

The material and methods of this study followed the criteria of the METRICS^35^ radiomic quality score that assesses the methodological quality of radiomic research and achieved a score of 84% (Figure S1).

## Results

### Patient Characteristics

A total of 380 patients (mean age, 65 ± 11 years; 218 males [57%]) were retrospectively included in our study, with patients excluded based on the exclusion criteria (Figure 1). Most patients had lung adenocarcinoma (279 [73%]), were current or former smoker (327 [86%]), were treated by chemotherapy (325 [86%]) and radiotherapy (247 [65%], Table 1). Depressive disorders were recorded in the medical records in 18/380 (5%) patients. Median follow-up duration was 16.8 months (IQR, 0.67-124.4 months). A total of 147 out of 380 (39%) patients died within 1 year.

**Table 1:** Demographic and Clinical Characteristics of Patients with metastatic NSCLC.

| Characteristics | Eligible Cohort<br>(n = 380) | Discovery set<br>(n = 234) | Test set<br>(n = 146) | P-Value* |
| --- | --- | --- | --- | --- |
| Age at diagnosis (y) | 65 ± 11 | 64 ± 11 | 66 ± 11 | .15 |
| Body Mass Index (kg/m<sup>2</sup>) | 23.8 ± 4.0 | 23.4 ± 4.0 | 24.4 ± 3.9 | .004 |
| Histology |  |  |  | .43 |
| Adenocarcinoma | 279 (73) | 167 (71) | 112 (77) |  |
| Squamous Cell Carcinoma | 52 (14) | 36 (15) | 16 (11) |  |
| Other | 49 (13) | 31 (14) | 18 (12) |  |
| Performance Status |  |  |  | .04 |
| 0-1 | 307 (81) | 181 (77) | 126 (86) |  |
| ≥2 | 73 (19) | 53 (23) | 20 (14) |  |
| Sex |  |  |  | .96 |
| Female | 162 (43) | 99 (42) | 63 (43) |  |
| Male | 218 (57) | 135 (58) | 83 (57) |  |
| Smoking history |  |  |  | .57 |
| Never | 53 (14) | 35 (15) | 18 (12) |  |
| Current or Former | 327 (86) | 199 (85) | 128 (88) |  |
| Depressive disorders |  |  |  | .84 |
| Absence/Not specified | 362 (95) | 222 (95) | 140 (96) |  |
| Presence | 18 (5) | 12 (5) | 6 (4) |  |
| Brain metastasis |  |  |  | .31 |
| Absence | 293 (77) | 185 (79) | 108 (74) |  |
| Presence | 87 (33) | 49 (21) | 38 (26) |  |
| Treatment received |  |  |  |  |
| Chemotherapy | 325 (86) | 200 (85) | 125 (86) | > .99 |
| Immunotherapy | 222 (58) | 106 (45) | 116 (79) | < .001 |
| Radiotherapy | 247 (65) | 154 (66) | 93 (64) | .76 |
| Targeted therapy | 122 (32) | 80 (34) | 42 (29) | .32 |
| 1-year survival |  |  |  | < .001 |
| Alive | 233 (61) | 131 (56) | 102 (70) |  |
| Dead | 147 (39) | 103 (44) | 44 (30) |  |
| Follow-up duration (months) | 24.6 ± 22.3 | 25.1 ± 25.5 | 23.7 ± 15.9 | .16 |

Note.—Categorical data are numbers of patients, with percentages in parentheses; continuous data are means, with standard deviation. \* The P-values from the Wilcoxon and $\chi^2$ tests between the discovery and test sets are shown.

Our study cohort was divided into 234 (62%) patients in the discovery set and 146 (38%) patients in the test set based on their scan date. Patients in the test set had higher BMI (*P* = .004), ECOG PS of 0-1 (126 [86%], *P* = .04) and were mostly treated with immunotherapy (116 [79%], *P* < .001) compared to patients in the discovery set.

### Evaluation of automatic segmentation

The tumor regions segmented by LION and those segmented by the experts were very similar, with an average DICE score of 0.90 ± 0.16 (range: [0.0; 1.0]) and no substantial differences in the derived radiomic feature values based on the Bland-Altman analyses (Figure S2). The prognostic performance of the five radiomic feature was similar for both segmentation approaches, except for Dmax (significant for the expert and non-significant for LION) (Figure S3). To ensure replicability, the results presented below were obtained using the fully automated LION segmentation refined with a threshold of 4 SUV.

### Survival Analysis

Among the 380 patients, SUVmean_brain_ showed a significant but weak negative correlation with TMTV (rS = -0.24, *P* < .001) and TTLG (rS = -0.21, *P* < .001) (Figure S4). There was no significant correlation (*P* > .05) between SUVmean_brain_ and Dmax (rS = -0.09), SUVmean_brain_ and TSUVmean (rS = 0.03) and SUVmean_brain_ and maxSUVmax (rS = 0.06). TTLG was excluded from the subsequent univariable and multivariable analyses due to its high significant correlation with TMTV (rS = 0.98, *P* < .001).

The Kaplan Meier curves for each feature are shown in Figure S5 (continuous variable dichotomized by the median). In the discovery set, these univariable analyses showed that five variables were statistically associated with OS (Table 2): ECOG (HR = 2.20, 95% CI 1.60-3.03, *P* < .001), smoking history (HR = 1.63, 95% CI 1.10-2.42, *P* = .01), Dmax (HR = 1.07, 95% CI 1.02-1.13, *P* = .009), TMTV (HR = 1.12, 95% CI 1.06-1.19, *P* < .001), and SUVmean_brain_ (HR = 0.83, 95% CI 0.76-0.92, *P* < .001). In the multivariable analysis without SUVmean_brain_ (Model 1), all these features were significantly associated with OS except Dmax: ECOG (HR = 2.04, 95% CI 1.47-2.82, *P* < .001), smoking history (HR = 1.58, 95% CI 1.06-2.35, *P* = .03) and TMTV (HR = 1.06, 95% CI 1.06-1.19, *P* < .001). Using Model 1, patients from the discovery set were divided into a high-risk and a low-risk groups based on the risk score calculated from the multivariable Cox model dichotomized by the median value (1.54 [IQR, 0.71–42.1]). OS was significantly different (*P* < .001) between the two risk groups as shown by the Kaplan Meier curves (Figure 2A-B). Median OS was 23.4 months (95% CI 17.4–30.9 months) and 9.0 months (95% CI 7.4-12.0 months) in the low-risk and high-risk groups respectively.

**Figure 2:**
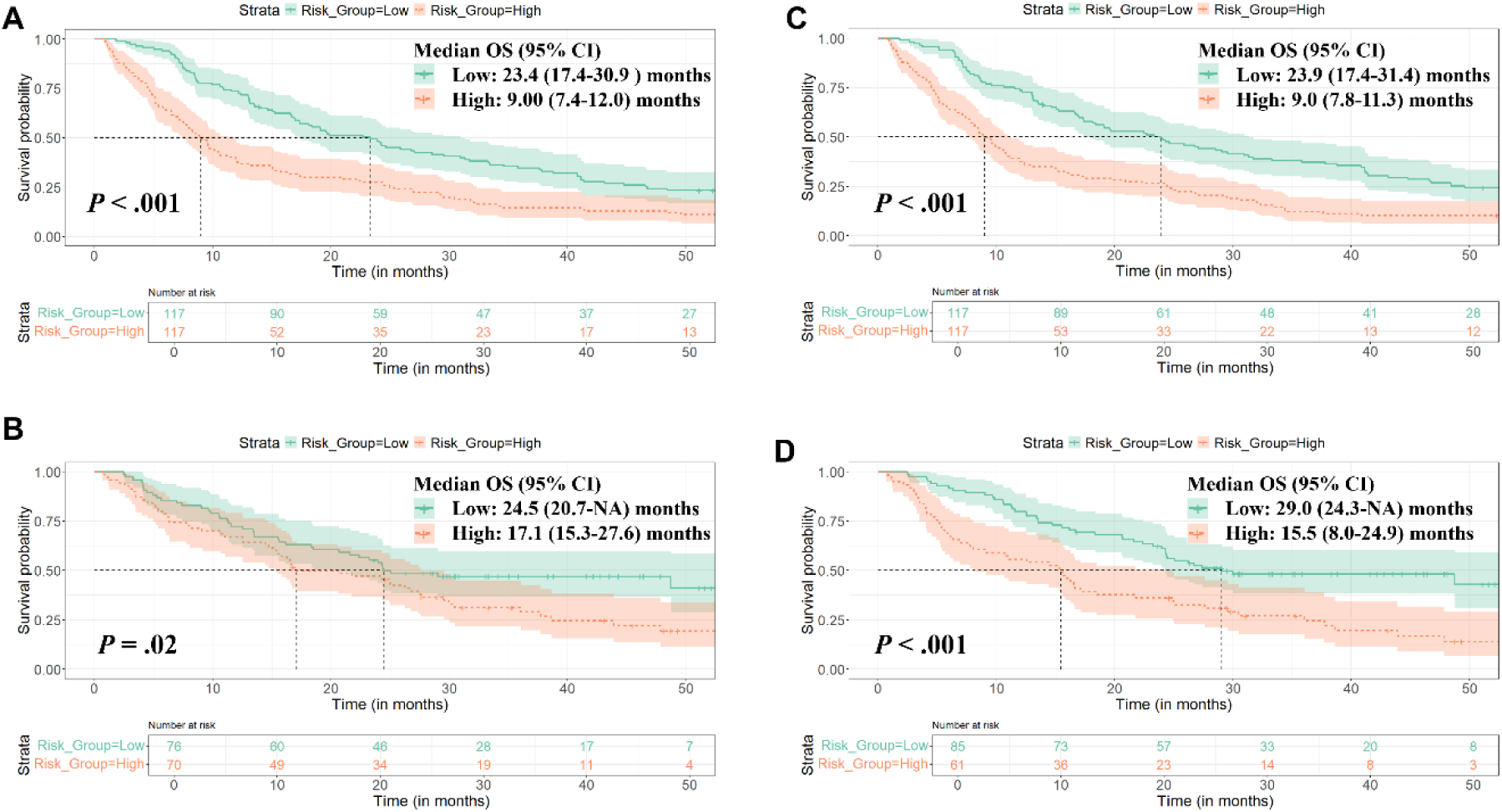
Kaplan-Meier curves for two multivariate models in the discovery (n = 234) and test sets (n = 146). **(A-B)** Model 1 including Performance Status, Smoking history, Dmax, TMTV and maxSUVmax for discovery **(A)** and test **(B)** sets. **(C-D)** Model 2 including features from Model 1 and SUVmean_brain_ for discovery **(C)** and test **(D)** sets. Patients were stratified into risk groups using the median value (1.54 for Model 1 and 1.43 for Model 2) derived from the discovery set. The Log-rank test *P*-values are shown.

**Table 2:** Cox Proportional Hazards Regression Assessing the Association of Clinical and Radiomic Features and SUVmean_brain_ with Overall Survival in the Discovery set (n = 234).

| Variable | Univariable analysis |  | Model 1 |  | Model 2 |  |
| --- | --- | --- | --- | --- | --- | --- |
|  | HR | P-Value | HR | P-Value | HR | P-Value |
| Age (years) | 1.01 (1.00-1.02) | .08 |  |  |  |  |
| BMI (kg/m<sup>2</sup>) | 0.99 (0.95-1.03) | .56 |  |  |  |  |
| Performance Status |  |  |  |  |  |  |
| 0-1 | - |  | - |  | - |  |
| ≥2 | 2.20 (1.60-3.03) | < .001 | 2.04 (1.47-2.82) | < .001 | 1.98 (1.43-2.74) | < .001 |
| Sex |  |  |  |  |  |  |
| Female | - |  |  |  |  |  |
| Male | 1.19 (0.90-1.57) | .21 |  |  |  |  |
| Smoking history |  |  |  |  |  |  |
| Never | - |  | - |  | - |  |
| Current or Former | 1.63 (1.10-2.42) | .01 | 1.58 (1.06-2.35) | .03 | 1.40 (0.93-2.12) | .11 |
| Dmax/10 | 1.07 (1.02-1.13) | .009 | 1.05 (0.99-1.10) | .10 | 1.04 (0.99-1.10) | .13 |
| TMTV/100 | 1.12 (1.06-1.19) | < .001 | 1.06 (1.06-1.19) | < .001 | 1.11 (1.04-1.17) | < .001 |
| TTLG/100 | 1.02 (1.01-1.02) | < .001 |  |  |  |  |
| TSUVmean | 1.05 (0.99-1.12) | .11 |  |  |  |  |
| maxSUVmax | 1.00 (1.00-1.01) | .06 |  |  |  |  |
| SUVmean<sub>brain</sub> | 0.83 (0.76-0.92) | < .001 |  |  | 0.88 (0.80-0.98) | .02 |

Note.— This table describes the association between clinical data, radiomic features and brain metabolic activity measured from PET, and mortality. Statistical analyses were performed using uni- and multivariable Cox proportional hazard regression models. Values in parentheses indicate the 95% confidence intervals). \*Model 1: all variables associated with OS at the $P < .05$ level in univariable analysis were included (Performance Status, Smoking history, Dmax, TMTV) except SUVmean<sub>brain</sub>; †Model 2: features from model 1 and SUVmean<sub>brain</sub>. The Wald test $P$ -values are shown.

When SUVmean_brain_ was used to build multivariable Model 2, a significantly better patient stratification (*P* = .02) was observed compared to Model 1, with a median survival of 23.9 months for low-risk patients and 9.0 months for high-risk patients (Log-rank test, *P* < .001), resulting a difference ΔOS of 14.9 months compared to 14.4 months with Model 1.

When Model 1 was applied to the test cohort, OS was also significantly different between the two risk groups (*P* = .02) with a median OS of 24.5 months (95% CI 20.7–NA months) for 76 low-risk patients and 17.1 months (95% CI 15.3–27.6 months) for 70 high-risk patients respectively. The addition of SUVmean_brain_ (Model 2) improved the distinction between low and high-risk patients (*P* < .001) in that test set, with a difference in median survival ΔOS between the two patient groups of 13.5 months, compared to 7.4 months with Model 1. Very similar results were obtained when using SULmean_brain_ (Table S2, Figure S6) instead of SUVmean_brain_.

### Identification of factors influencing cerebral metabolic activity

Among the 87 (23%) patients with brain metastases, only 29 (33%) had either metastases or perilesional oedema visible on the PET images. Low value of cerebral metabolic activity was significantly associated with poor OS (*P* < .001), regardless of whether based on VOI_brain or VOI_brain_woM (Figure 3A, Figure S7 A-C). When the whole-brain was automatically segmented by TotalSegmentator (VOI_brain), we observed no significant difference in SUVmean_brain_ distribution between patients with metastases in the brain and those without (*P* = .68). Patients who died within the first year of follow-up (233/380 patients [61%]) had a significantly lower SUVmean_brain_ value than patients who were alive after 1 year (*P* < .001, Figure 3B, Figure S7 B-D). Similar results were found for SULmean_brain_ (Figure S8). When stratifying patients based on the treatment received (Figure S9), the prognostic value of SUVmean_brain_ was confirmed for patients who received chemotherapy (235/380 patients [86%], *P* < .001), radiotherapy (247/380 patients [65%], *P* = .01), or immunotherapy (222/380 patients [58%], *P* = .005), but did not reach statistical significance in patients treated with targeted therapies (122/380 patients [32%], *P* = .16).

**Figure 3:**
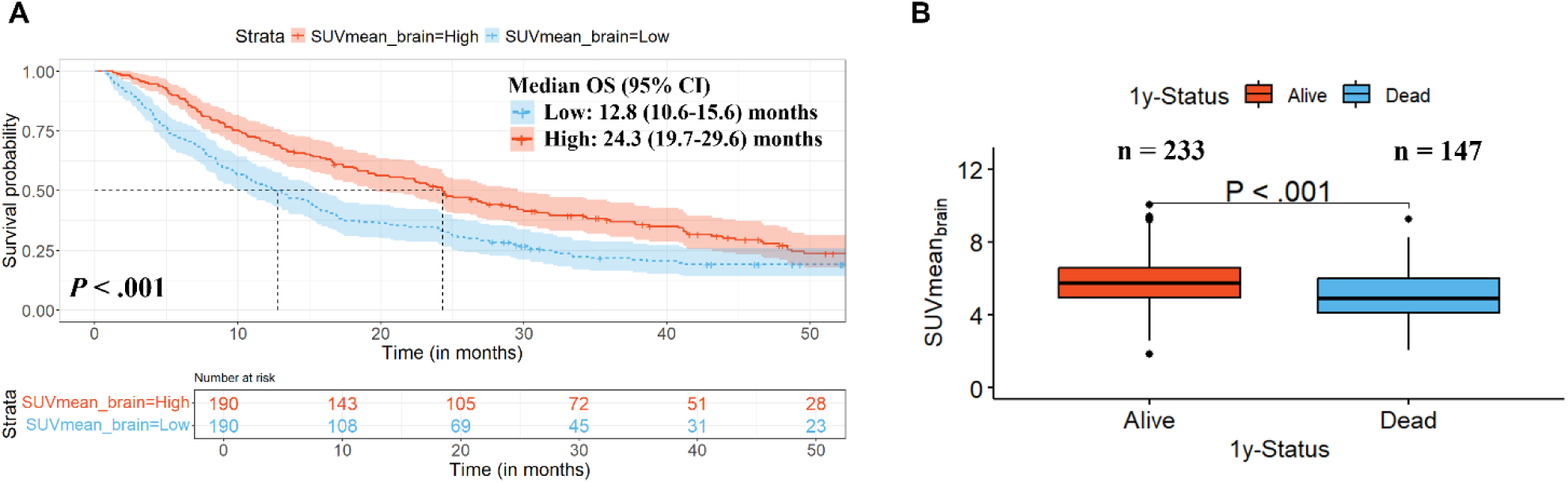
Relationship between SUVmean_brain_ and overall survival in metastatic NSCLC patients. **(A)** Kaplan Meier curves for NSCLC patients in the Eligible Cohort (n = 380) stratified into risk groups by the median of the SUVmean_brain_ (median = 5.45). Log-rank test *P*-value is shown. **(B)** Boxplot representation of SUVmean_brain_ according to the 1-year vital status in the Eligible Cohort. Wilcoxon test *P*-value is shown.

SUVmean_brain_ was significantly but weakly correlated with BMI (rS = 0.26, Figure S10) and negatively correlated with the patient’s age (rS = -0.20). Moreover, SUVmean_brain_ was significantly lower in current/smoker patients than in non-smoker patients (*P* = .002) as well as in PS ≥ 2 compared to PS = 0-1 patients (*P* < .001). There was no significant correlation between SUVmean_brain_ and the technical parameters influencing uptake like the injected dose, the time from injection to acquisition and the acquisition duration (Figure S11A).

Regarding biological data, blood biomarkers were available for 110/380 patients (29%) (Figure S11B, Figure S12). SUVmean_brain_ was positively correlated with albumin (rS = 0.31, *P* < .001), lymphocytes (rS = 0.15, *P* = .02), PNI (rS = 0.35, *P* < .001), and negatively correlated with blood glucose levels (rS = -0.43, *P* < .001), neutrophils (rS = -0.38, *P* < .001), CRP (rS = -0.37, *P* < .001), leukocytes (rS = -0.37, *P* < .001) and monocytes (rS = -0.32, *P* < .001). There was a slight but non-significant correlation (*P* = .26) between SUVmean_brain_ and cortisol (rS = -0.18). Kaplan Meier curves of each blood parameter enabling patient stratification into two groups of risk according to the normal biological values are shown in Figure S13 A-F. Patients who died within the first year of follow-up (46/110 patients [43%]) had higher value of CRP, leukocytes, monocytes and neutrophils and lower values of lymphocytes compared with patients who were still alive after 1 year (P ≤ .03, Figure S13 G-L). However, there were no significant differences in albumin levels between patients who were alive after 1 year and those who were not (*P* = .30). Blood glucose levels was significantly associated with survival (*P* < .001, Figure S14), with higher values observed in patients who died within the first year of follow-up (*P* = .02). The association of SUVmean_brain_ with blood biomarkers is shown in Figure S15 and Figure S16. Kaplan–Meier curves showed significantly different OS among patients stratified by risk factors (*P* ≤ .04), except for cortisol (*P* = .42, data available for only 42 patients), supporting the complementarity of the prognostic values of SUVmean_brain_ and blood biomarkers. Although low PNI was significantly associated with poorer survival (*P* = .02), its combination with SUVmean_brain_ did not provide additional prognostic value (Figure S17).

Finally, patients with known depressive disorders (n = 18) had lower brain FDG uptake (median SUVmean_brain_ = 4.94 [IQR, 2.45-7.76] and SULmean_brain_ = 3.87 [2.11-5.84]) than those without (SUVmean_brain_ = 5.48 [1.87-10.0] and SULmean_brain_ = 4.24 [1.52-7.42]), but the difference was not statistically significant (*P* = .33 and *P* = .24 respectively, Figure S18). Having depressive disorders, collected retrospectively from the medical records, was not significantly associated with OS.

Examples of patients with similar clinical and radiomic features but different SUVmean_brain_ levels are shown in Figure 4.

**Figure 4:**
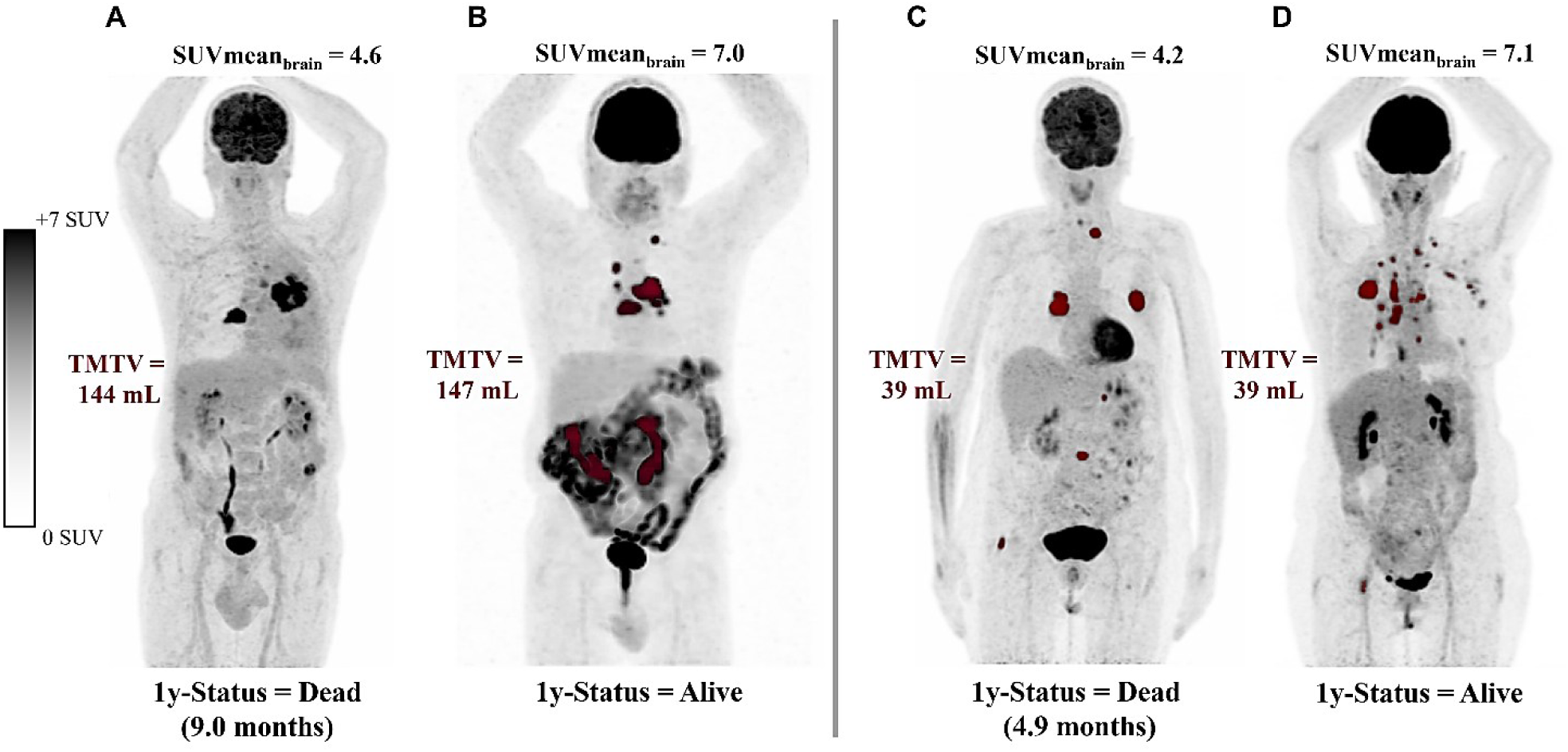
Examples of baseline 18F[FDG] PET images of four patients with metastatic NSCLC. Primary tumor and metastatic lesions are delineated in red. TMTV, SUVmean_brain_ values and 1-year survival status are shown. **(A)** A male in his 60s with a BMI of 22.4 kg/m², PS 0-1, smoker, blood glucose levels of 13.0 mmol/L. **(B)** A male in his 60s with a BMI of 24.8 kg/m², PS 0-1, smoker, blood glucose levels of 4.7 mmol/L. **(C)** A female in her 50s with a BMI of 23.1 kg/m², PS 0-1, smoker, blood glucose levels of 6.33 mmol/L. **(D)** A female in her 50s with a BMI of 23.2 kg/m², PS 0-1, smoker, blood glucose levels of 7.1 mmol/L.

## Discussion

Using whole-body [18F]FDG-PET/CT images, recent studies suggested that metabolic activity measured across various organs and tissues outside the tumor site could provide valuable information for predicting patients’ outcomes in lung cancer^11–14^. Our main finding was that brain FDG uptake was an independent prognostic factor in both univariable (HR = 0.83, *P* < .001) and multivariable analyses (HR = 0.88, *P* = .02) according to Cox models and complemented radiomic features reflecting tumor invasion. Brain FDG uptake combined with clinical and tumor-related radiomic features measured from baseline [18F]-FDG-PET/CT scans stratified metastatic NSCLC patients into low-risk and high-risk groups according to overall survival (*P* < .001). In the discovery set (n = 234), median OS was 23.9 months in low-risk and 9.0 months in high-risk groups. In the test set (n = 159), median OS was 29.0 months in low-risk and 15.5 months in high-risk groups. In the whole cohort, patients who died within one year of follow-up (173/380 [38%]) had significantly lower SUVmean_brain_ (*P* < .001) than those with a life expectancy > 1y (280/380 [62%]). Low brain FDG uptake was always significantly associated with a poor prognosis, regardless of the treatment received after PET/CT (*P* ≤ .01), except for targeted therapy (*P* = .16 with only 122 patients in this treatment group). To ensure robust segmentation and reproducible results, all segmentations were performed using fully-automated tools (LION and TotalSegmentator), which reduces operator dependency and saves time. In particular, we demonstrated a very good agreement between manual segmentation by an expert and the segmentation performed by LION in metastatic patients.

Several assumptions can be made to explain our observations. One is a possible relationship between brain metabolism and the systemic inflammation. In our study, low brain FDG uptake was associated with poorer overall survival than high brain FDG uptake (*P* < .001, median OS 12.8 vs 24.3 months). We observed moderate but significant correlations between SUVmean_brain_ and peripheral blood biomarkers, such as albumin levels, CRP, Blood glucose levels leukocyte, lymphocyte, monocyte and neutrophil counts (|rS| range: 15 to 43, *P* ≤ .02). Numerous studies have shown that blood markers evaluated individually or combined into scores were significant prognostic factors for overall survival in patients with lung cancer^18–22^. Moreover, combining blood-based biomarkers with PET-derived biomarkers such as TMTV has been suggested to provide a more integrated prognostic approach in NSCLC^36^. We demonstrated that high values of CRP, leukocytes, monocytes, neutrophils and blood glucose levels were significantly associated with a poor OS (*P* ≤ .02). The combination of these blood biomarkers with SUVmean_brain_ resulted in the stratification of patients into three different groups with significantly different overall survival (*P* ≤ .04), except for cortisol (*P* = .42) possibly due to the limited sample size (n = 42). Therefore, these findings support the complementary prognostic value of SUVmean_brain_ and peripheral blood biomarkers and highlight the relevance of their combination for the stratification of metastatic NSCLC patients according to OS.

Another possible explanation could be a correlation between cerebral FDG uptake and the patient’s functional status, encompassing both physical and cognitive components. Our analysis demonstrates that cerebral FDG uptake is weakly but significantly correlated with several factors, including age, BMI, sex, smoking status, ECOG PS, serum albumin levels, and blood glucose levels, in line with previous studies that have individually reported associations between some of these parameters and cerebral uptake^37–40^. Taken together, these findings suggest that SUVmean_brain_ may reflect the patient’s overall functional status, including ability to function and nutritional status, which are known to be associated with clinical prognosis^41–42^. Other studies using [18F]-FDG-PET/CT imaging have reported a correlation between decreased cerebral glucose metabolism in specific brain regions and patient’s psychological conditions, with or without cancer^43–46^. In oncology, patients face a higher risk of depression, with a prevalence influenced by various factors, including sex, age, and cancer type^47^. It has been reported that patients with lung cancer had lower brain FDG uptake than in patients with other cancer types, and a higher incidence of depression and anxiety, particularly in metastatic patients^45^. In our study, patients with known depressive disorders (18/380 [5%]) had lower brain FDG uptake than those without (362/380 [95%]), however the difference was not statistically significant (*P* = .33). Moreover, several studies have shown that there is a link between patients with depressive syndromes and blood biomarkers indicating unfavorable systemic inflammation^48–51^ and between nutritional status and mental condition^52^. In our study, SUVmean_brain_ was positively correlated with PNI (rS = 0.35, *P* < .001); however, their combination did not improve prognostic stratification according to OS. Further studies are now needed to elucidate the biological mechanisms reflected by the brain metabolism.

A limitation of our study was that there were significant differences between the composition of the discovery and test sets. We divided the cohort based on the PET scan date, which introduced a bias in the treatment selection and overall survival. Indeed, patients in the test cohort had access to more innovative treatments such as immunotherapy. This results in a difference in survival between the discovery and the test cohorts. Nevertheless, the models could still stratify the patients of the test set without adjusting the cut-off values, although the used cut-off value is likely to be sub-optimal. Another limitation was the incomplete availability of biological data for some patients, which reduced the cohort sizes and may have limited the statistical power of some analyses. Finally, we collected information retrospectively on depression from patient medical records, which is certainly incomplete and imperfect as patient may not necessarily report a depressive disorder or may be unaware of it. To demonstrate a link between SUVmean_brain_, patient’s functional status and survival, we would need to prospectively collect a score measuring depression at the time of the baseline [18F]-FDG PET/CT scan such as the Hamilton Depression Rating Scale^53–60^, a score assessing cognitive abilities, such as Mattis Dementia Rating Scale 2, and also use connected devices to measure the patient’s physical condition and activity. Further studies should be conducted to test the hypothesis regarding the relationship between cerebral FDG uptake and the patient’s functional status. Depending on the results of these investigations, SUVmean_brain_ could potentially be used to identify patients who should be prioritized for supportive care interventions such as psychological support, nutritional counselling, or tailored physical activity programs.

In conclusion, our study showed that low brain FDG uptake was associated with increased risk of mortality in patients with metastatic NSCLC. Furthermore, our prognostic model, which combined brain FDG uptake, clinical and tumor-related radiomic features, improved overall survival prediction compared to a model based only on clinical and tumor-related radiomic features. Further studies investigating the role of brain FDG uptake as a prognostic biomarker for overall survival and its association with systemic inflammation and the patient’s functional status are warranted.

## Data Availability

All data produced in the present study are available upon reasonable request to the authors.

## ACKNOWLEDGEMENTS

This work was supported by the French National Research Agency: ANR-22-CE45-0001 NEMO-PET. We would like to thank the following collaborators from Data Office of Institut Curie for their valuable support: Maud Milder and Laetitia Chanas.

