## Supplemental data for "Brain Metabolic Activity on FDG PET/CT Predicts Survival in Non–Small Cell Lung Cancer"

#### Supplemental Materials

**Table S1: Imaging protocol parameters.**

| Scanner model | Manufacturer name | Slice thickness (mm) | Pixel spacing (mm) | Reconstruction method |
|---|---|---|---|---|
| Discovery 610 | GE | 3.27 | [2.73; 2.73] | VPHDS |
| Discovery 690 | GE | 3.27 | [2.73; 2.73] | VPFXS, VPFX |
| Discovery 710 | GE | 3.27 | [2.73; 2.73] | VPFXS, VPFX |
| Discovery IQ | GE | 3.26 | [2.73; 2.73] | QCHD |
| Discovery MI | GE | 2.79 | [1.82; 1.82] | QCFX |
| Discovery MI | GE | 2.79 | [2.73; 2.73] | VPFXS, QCFX |
| Discovery RX | GE | 3.27 | [5.47; 5.47] | 3D IR |
| Discovery ST | GE | 3.27 | [2.73; 2.73] | 3D IR |
| Discovery STE | GE | 3.27 | [5.47; 5.47] | 3D IR |
| Optima 560 | GE | 3.27 | [2.73; 2.73] | VPHD |
| GEMINI TF TOF 16 | Philips | 2.00 | [2.00; 2.00] | BLOB-OS-TF |
| GEMINI TF TOF 16 | Philips | 4.00 | [4.00; 4.00] | BLOB-OS-TF |
| GEMINI TF TOF 64 | Philips | 2.00 | [2.00; 2.00] | BLOB-OS-TF |
| Ingenuity TF PET/CT | Philips | 2.00 | [2.00; 2.00] | BLOB-OS-TF |
| Biograph 20_mCT | SIEMENS | 2.03 | [4.07; 4.07] | PSF+TOF 2i21s |
| Biograph 20_mCT | SIEMENS | 5.00 | [4.07; 4.07] | PSF+TOF 3i21s |
| Biograph 40_mCT | SIEMENS | 2.03 | [4.07; 4.07] | PSF 4i12s |
| Biograph 40_mCT | SIEMENS | 2.03 | [4.07; 4.07] | PSF+TOF 2i21s |
| Biograph 40_mCT | SIEMENS | 4.00 | [4.07; 4.07] | OSEM3D+TOF 2i21s |
| Biograph 40_mCT | SIEMENS | 5.00 | [4.07; 4.07] | PSF+TOF 2i21s |
| Biograph Horizon | SIEMENS | 2.03 | [2.06; 2.06] | PSF+TOF 6i10s |
| Biograph6_True Point | SIEMENS | 5.00 | [4.07; 4.07] | OSEM2D 6i8s |
| Biograph64_mCT | SIEMENS | 2.03 | [4.07; 4.07] | PSF 4i12s |
| Biograph64_Vision 450 | SIEMENS | 1.65 | [1.65; 1.65] | PSF+TOF 3i5s |
| Biograph64_Vision 450 | SIEMENS | 1.65 | [1.65; 1.65] | PSF+TOF 4i5s |
| Biograph64_Vision 600 | SIEMENS | 1.65 | [1.65; 1.65] | PSF+TOF 3i5s |

### METRICS Tool v1.0

**Please fill out all conditions first for relevant sections and then all active items to calculate METRICS score.**

**Please note that default option is "No".**

? Stands for explanation of items and conditions.

C Stands for conditional items or sections.

| Items/Conditions | Definitions | Weights | Options |
|---|---|---|---|
| **Study Design** | | | |
| Item#1 | ? Adherence to radiomics and/or machine learning-specific checklists or guidelines | 0.0368 | ◉ Yes ○ No |
| Item#2 | ? Eligibility criteria that describe a representative study population | 0.0735 | ◉ Yes ○ No |
| Item#3 | ? High-quality reference standard with a clear definition | 0.0919 | ◉ Yes ○ No |
| **Imaging Data** | | | |
| Item#4 | ? Multi-center | 0.0438 | ○ Yes ◉ No |
| Item#5 | ? Clinical translatability of the imaging data source for radiomics analysis | 0.0292 | ◉ Yes ○ No |
| Item#6 | ? Imaging protocol with acquisition parameters | 0.0438 | ◉ Yes ○ No |
| Item#7 | ? The interval between imaging used and reference standard | 0.0292 | ◉ Yes ○ No |
| **Segmentation** C | | | |
| Condition#1 | ? Does the study include segmentation? | | ◉ Yes ○ No |
| Condition#2 | ? Does the study include fully automated segmentation? | | ◉ Yes ○ No |
| Item#8 | ? Transparent description of segmentation methodology | 0.0337 | ◉ Yes ○ No |
| Item#9 | ? Formal evaluation of fully automated segmentation C | 0.0225 | ◉ Yes ○ No |
| Item#10 | ? Test set segmentation masks produced by a single reader or automated tool | 0.0112 | ◉ Yes ○ No |
| **Image Processing and Feature Extraction** | | | |
| Condition#3 | ? Does the study include hand-crafted feature extraction? | | ◉ Yes ○ No |
| Item#11 | ? Appropriate use of image preprocessing techniques with transparent description | 0.0622 | ◉ Yes ○ No |
| Item#12 | ? Use of standardized feature extraction software C | 0.0311 | ◉ Yes ○ No |
| Item#13 | ? Transparent reporting of feature extraction parameters, otherwise providing a default configuration statement | 0.0415 | ◉ Yes ○ No |
| **Feature Processing** | | | |
| Condition#4 | ? Does the study include tabular data? | | ◉ Yes ○ No |
| Condition#5 | ? Does the study include end-to-end deep learning? | | ○ Yes ◉ No |
| Item#14 | ? Removal of non-robust features C | 0.0200 | ○ Yes ◉ No |
| Item#15 | ? Removal of redundant features C | 0.0200 | ◉ Yes ○ No |
| Item#16 | ? Appropriateness of dimensionality compared to data size C | 0.0300 | ◉ Yes ○ No |
| Item#17 | ? Robustness assessment of end-to-end deep learning pipelines C | 0.0200 | ○ Yes ○ No |
| **Preparation for Modeling** | | | |
| Item#18 | ? Proper data partitioning process | 0.0599 | ◉ Yes ○ No |
| Item#19 | ? Handling of confounding factors | 0.0300 | ◉ Yes ○ No |
| **Metrics and Comparison** | | | |
| Item#20 | ? Use of appropriate performance evaluation metrics for task | 0.0352 | ◉ Yes ○ No |
| Item#21 | ? Consideration of uncertainty | 0.0234 | ◉ Yes ○ No |
| Item#22 | ? Calibration assessment | 0.0176 | ◉ Yes ○ No |
| Item#23 | ? Use of uni-parametric imaging or proof of its inferiority | 0.0117 | ◉ Yes ○ No |

| | | | | |
|---|---|---|---|---|
| Item#24 | ? Comparison with a non-radiomic approach or proof of added clinical value | 0.0293 | ◉ Yes | ○ No |
| Item#25 | ? Comparison with simple or classical statistical models | 0.0176 | ◉ Yes | ○ No |
| **Testing** | | | | |
| Item#26 | ? Internal testing | 0.0375 | ◉ Yes | ○ No |
| Item#27 | ? External testing | 0.0749 | ○ Yes | ◉ No |
| **Open Science** | | | | |
| Item#28 | ? Data availability | 0.0075 | ○ Yes | ◉ No |
| Item#29 | ? Code availability | 0.0075 | ○ Yes | ◉ No |
| Item#30 | ? Model availability | 0.0075 | ◉ Yes | ○ No |
| | | **Total METRICS score:** | **84.3%** | |
| | | ? **Quality category:** | **Excellent** | |
| | | ? **Publication ID:** | | |

**If you publish any work which uses this tool, please cite the following publication:**

Kocak B, Akinci D'Antonoli T, Mercaldo N, et al. METhodological RadiomICs Score (METRICS): a quality scoring tool for radiomics research endorsed by EuSoMII. Insights Imaging. 2024;15(1):8. Published 2024 Jan 17. doi:10.1186/s13244-023-01572-w

**Figure S1**: Evaluation of the quality of our study using the METRICS Score.

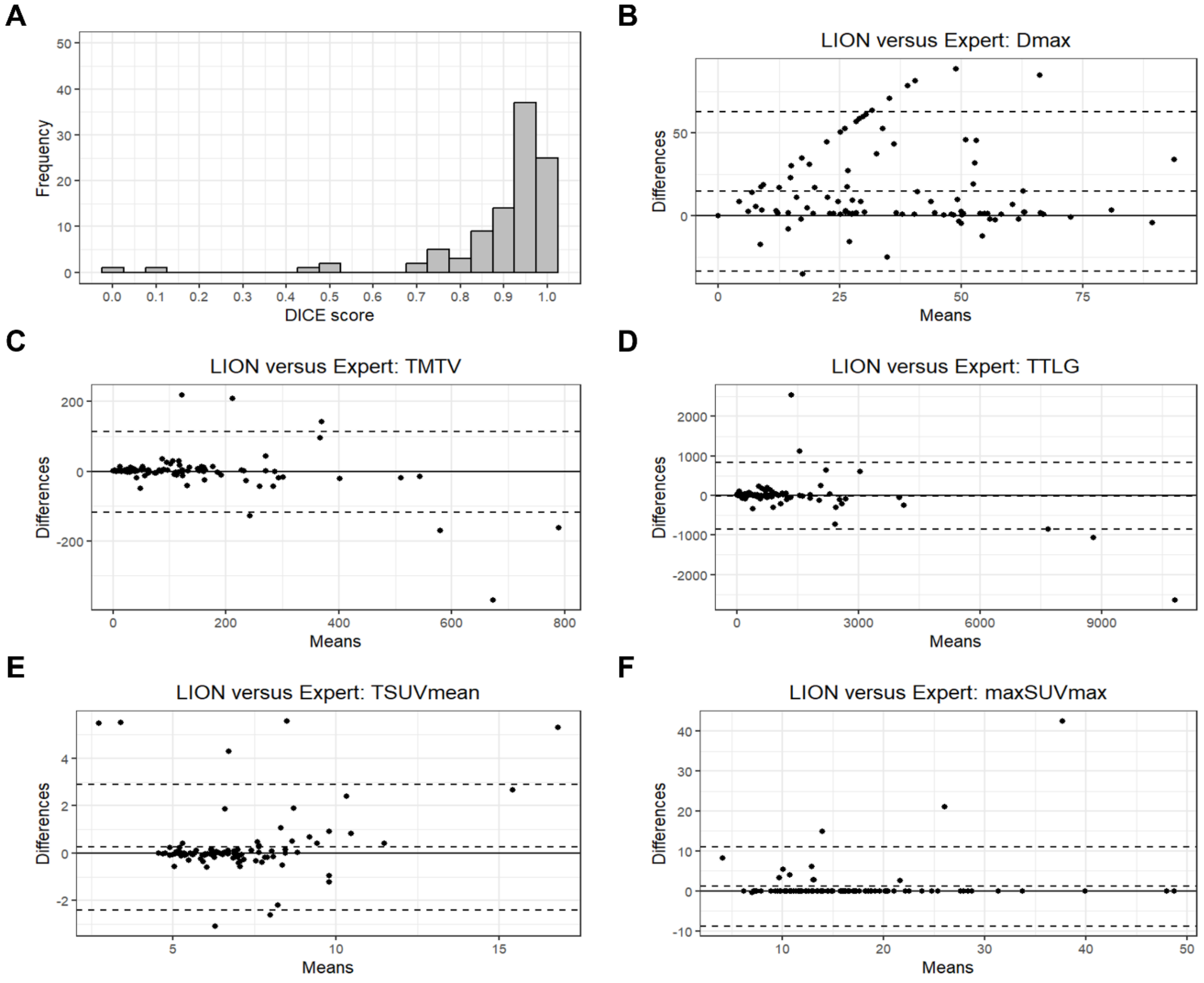


**Figure S2**: Comparison of lesion segmentation performance between LION algorithm and a 10-year expert for 100 NSCLC metastatic patients. **(A)** Histogram of Dice scores for the 100 patients, calculated by comparing the expert-segmented image with the LION-segmented image refined using a threshold of 4 SUV. Bland-Altman plots showing means on the x-axis and differences on the y-axis are presented for five standard radiomic features: **(B)** Dmax, **(C)** TMTV, **(D)** TTLG, **(E)** TSUVmean and **(F)** maxSUVmax.

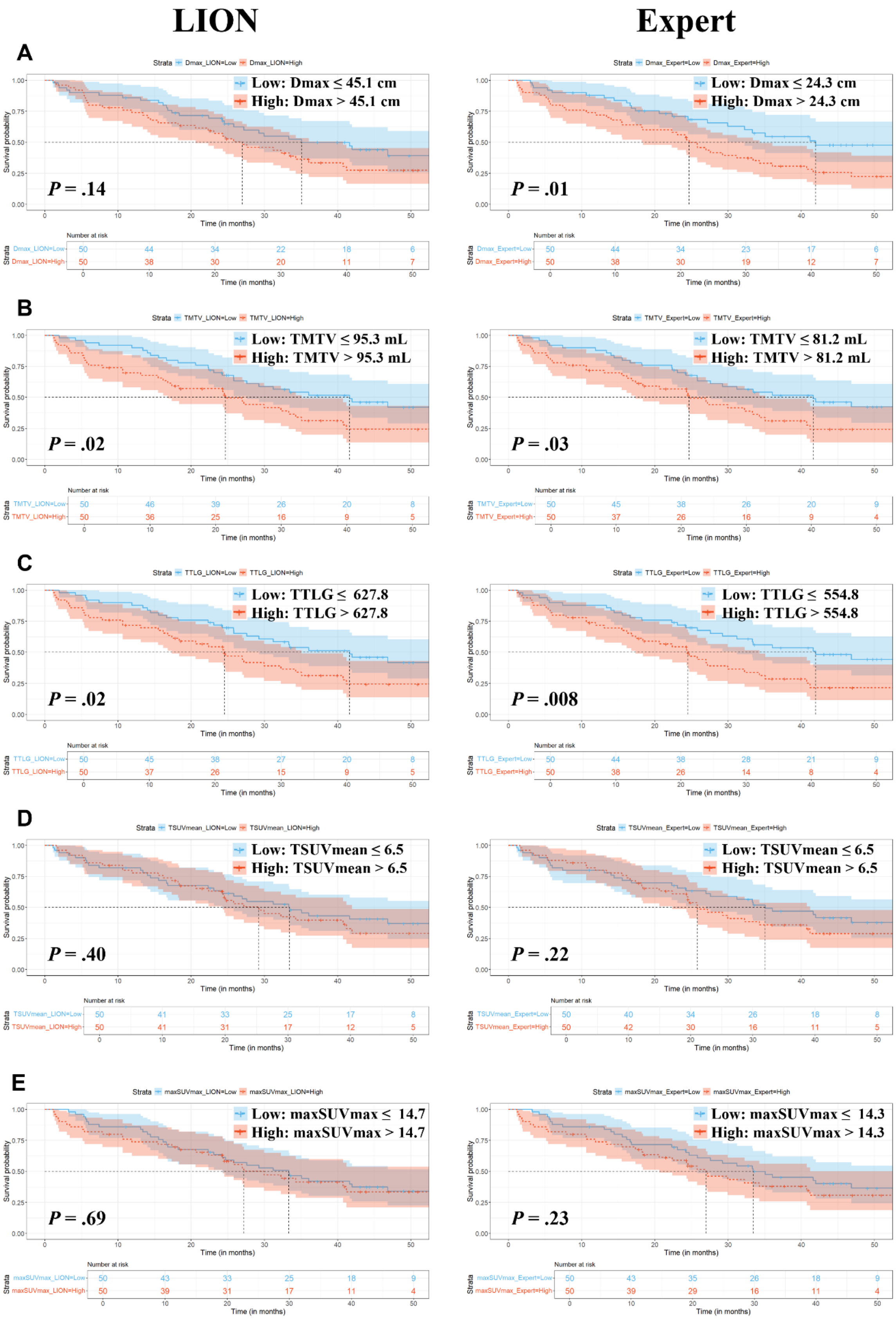


**Figure S3**: Kaplan-Meier curves of five radiomic features calculated from lesion segmentation produced by the LION algorithm and a 10-year expert in 100 NSCLC metastatic patients. Patients were stratified into high and low-risk groups according to OS based on features median which were calculated from LION algorithm (left) and from the expert (right) for Dmax **(A)**, TMTV **(B)**, TTLG **(C)**, TSUVmean **(D)** and maxSUVmax **(E)**. The Log-rank test *P* values are shown.

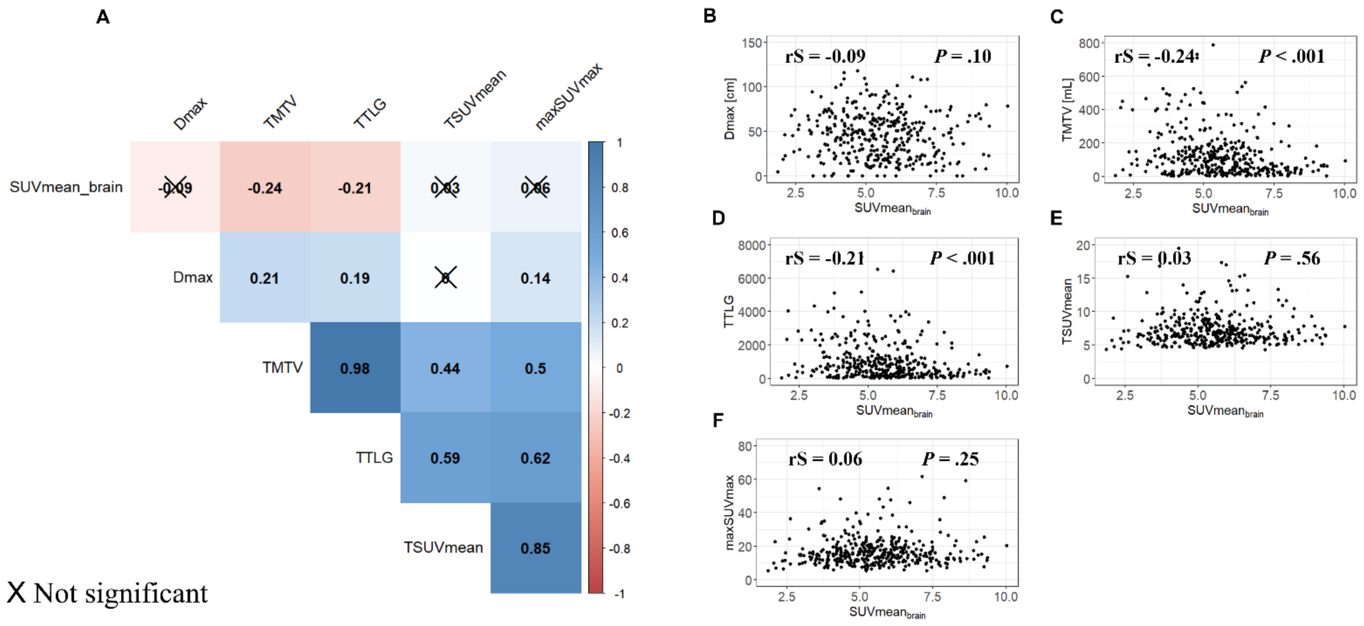


**Figure S4**: Relationship between $SUVmean_{brain}$ and radiomic features using Spearman correlation for in the full Cohort (n = 380). Spearman correlation coefficients (rS) and Wilcoxon test *P* values are shown. **(A)** Correlogram representation. **(B-F)** Correlation plot between $SUVmean_{brain}$ and Dmax **(B)**, TMTV **(C)** TTLG **(D)**, TSUVmean **(E)** and maxSUVmax **(F)**.

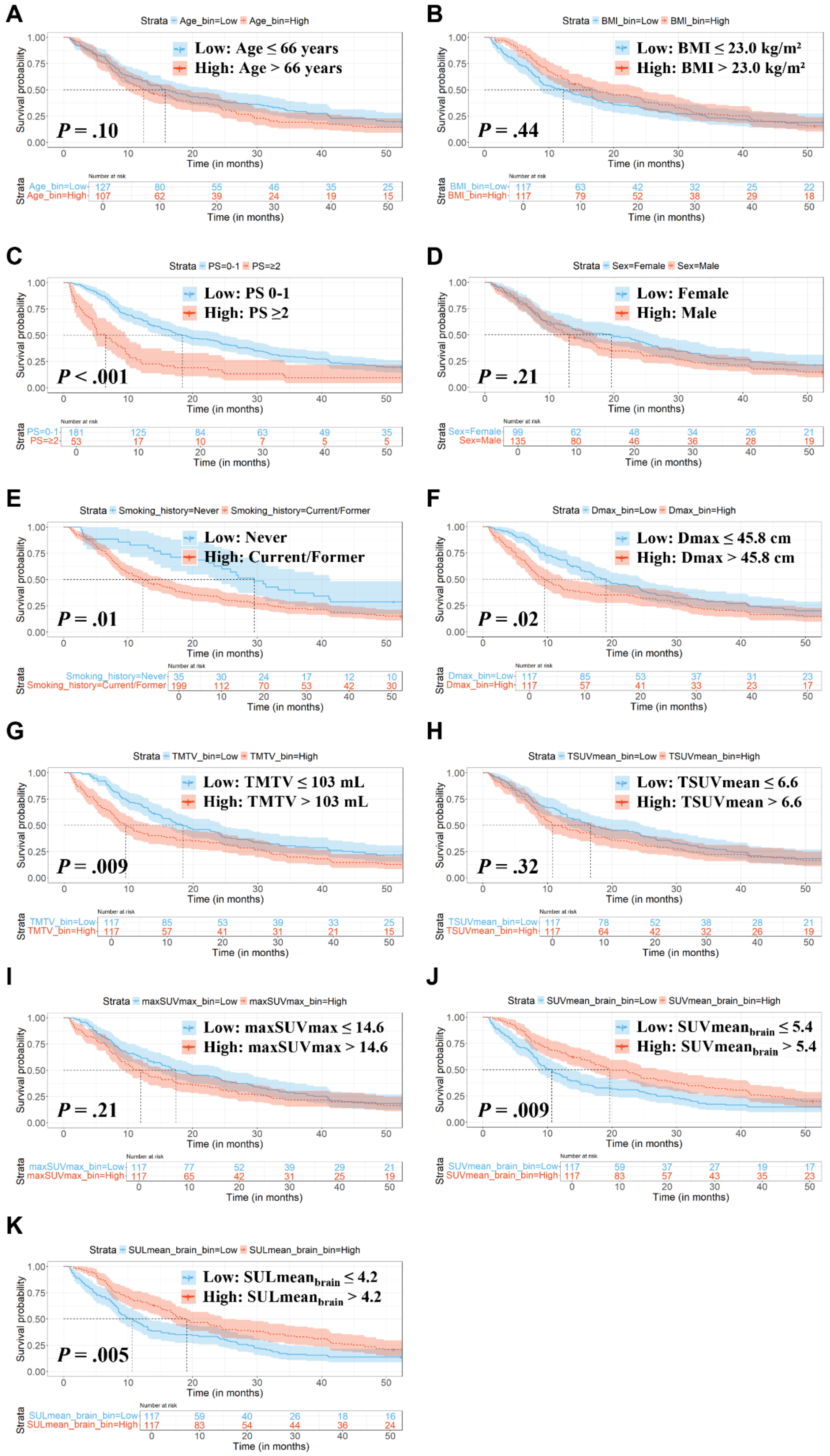


**Figure S5**: Kaplan-Meier curves of clinical and radiomic features used in univariable analysis according to OS for the discovery cohort (n = 234). **(A-K)** Patients are stratified into high and low-risk groups according to OS based on the median of feature values for quantitative data and labels for qualitative data. The Log-rank test *P* values are shown.

**Table S2: Cox Proportional Hazards Regression Assessing the Association of Clinical and Radiomic Features and $SULmean_{brain}$ with Overall Survival in the Discovery set (n = 234).**

| | Univariable analysis | | Model 1* | | Model 2† | |
|---|---|---|---|---|---|---|
| **Variable** | HR | *P* Value | HR | *P* Value | HR | *P* Value |
| **Age (years)** | 1.01 (1.00-1.02) | .08 | | | | |
| **BMI (kg/m²)** | 0.99 (0.95-1.03) | .56 | | | | |
| **Performance Status** | | | | | | |
| **0-1** | - | | - | | | |
| **≥2** | 2.20 (1.60-3.03) | < .001 | 2.04 (1.47-2.82) | < .001 | 1.98 (1.43-2.74) | < .001 |
| **Sex** | | | | | | |
| **Female** | - | | | | | |
| **Male** | 1.19 (0.90-1.57) | .21 | | | | |
| **Smoking history** | | | | | | |
| **Never** | - | | - | | | |
| **Current or Former** | 1.63 (1.10-2.42) | .01 | 1.58 (1.06-2.35) | .03 | 1.44 (0.96-2.16) | .08 |
| **Dmax/10** | 1.07 (1.02-1.13) | .009 | 1.05 (0.99-1.10) | .10 | 1.04 (0.99-1.10) | .11 |
| **TMTV/100** | 1.12 (1.06-1.19) | < .001 | 1.06 (1.06-1.19) | < .001 | 0.11 (1.04-1.17) | < .001 |
| **TSUVmean** | 1.05 (0.99-1.12) | .11 | | | | |
| **maxSUVmax** | 1.00 (1.00-1.01) | .06 | | | | |
| **$SULmean_{brain}$** | 0.79 (0.69-0.90) | < .001 | | | 0.74 (0.74-0.97) | .02 |

Note.— This table describes the association between clinical data, radiomic features and brain metabolic activity measured from PET, and mortality. Statistical analyses were performed using uni- and multivariable Cox proportional hazard regression models. Values in parentheses indicate the 95% confidence intervals (CIs). *Model 1: all variables associated with OS at the $P < .05$ level in univariable analysis were included (Performance Status, Smoking history, Dmax, TMTV, maxSUmax) except $SULmean_{brain}$; †Model 2: features from model 1 plus $SULmean_{brain.}$ The Wald test *P* values are shown.

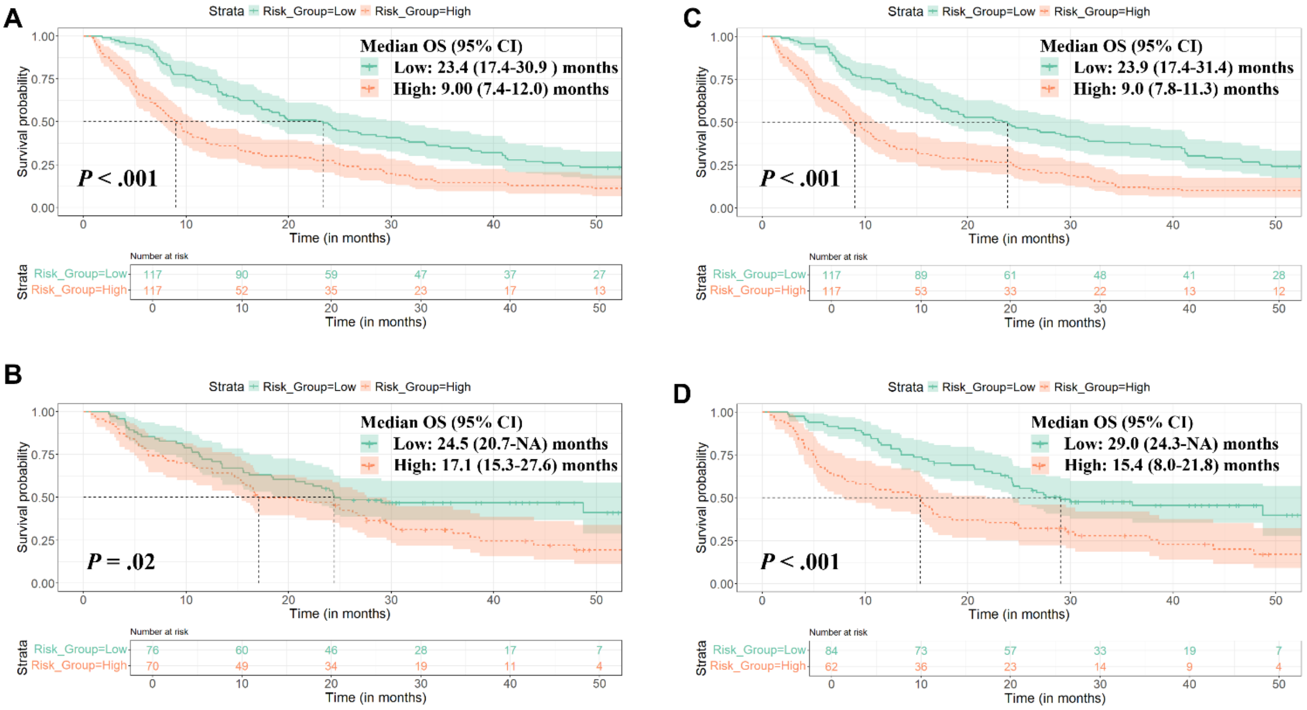


**Figure S6**: Kaplan-Meier curves for two multivariate models in the discovery (n = 234) and test sets (n = 146). **(A-B)** Model 1 including Performance Status, Smoking history, Dmax, TMTV and maxSUVmax for discovery **(A)** and test **(B)** sets. **(C-D)** Model 2 including features from model 1 and $SULmean_{brain}$ for discovery **(C)** and test **(D)** sets. Patients were stratified into risk groups using the median value derived from the discovery set (1.54 for model 1 and 1.44 for model 2). The Log-rank test *P* values are shown.

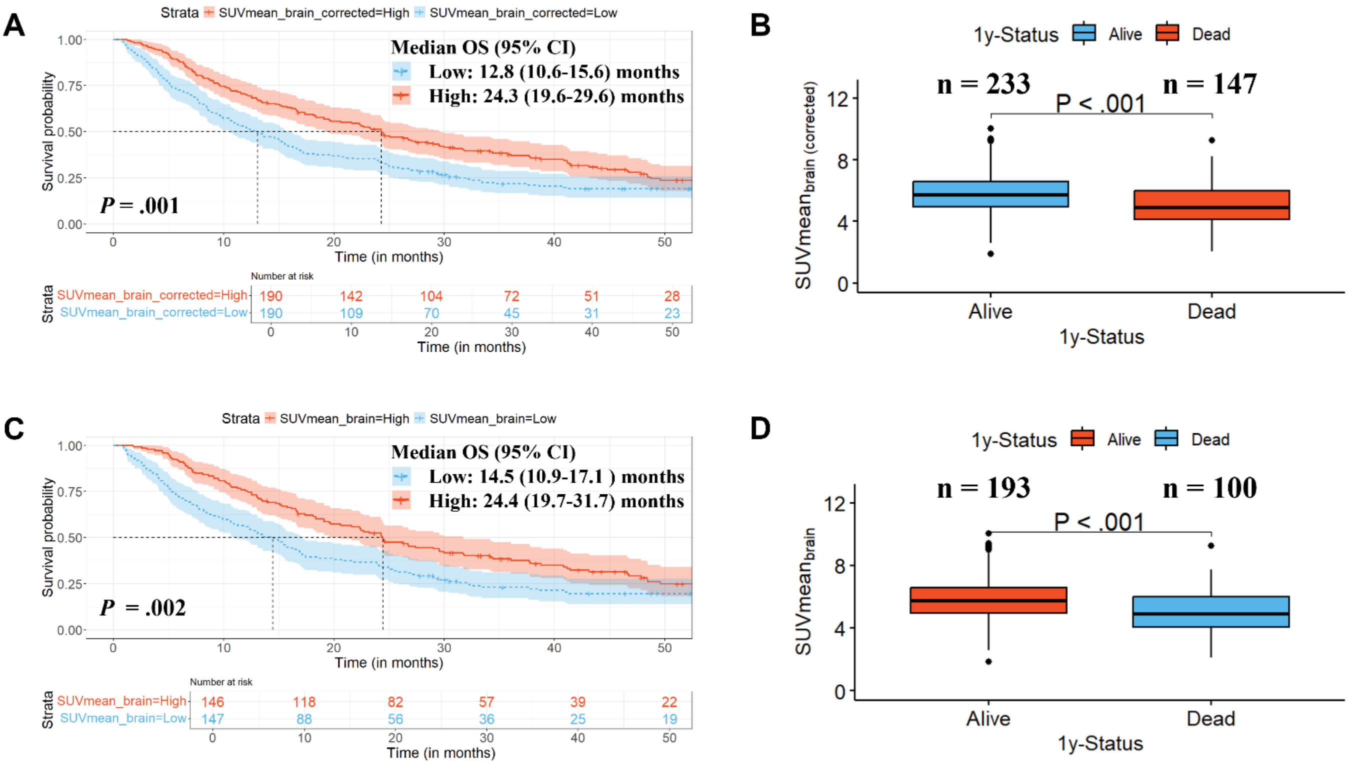


**Figure S7**: Relationship between brain metabolism, brain metastases and overall survival in metastatic NSCLC patients. (**A-C**) Kaplan Meier curves for NSCLC patients divided into 2 risk groups using the median of SUVmean$_{brain_noM}$ with brain metastases removed manually in the full cohort as a cut-off (n = 380, median = 5.46) **(A)** and SUVmean$_{brain}$ measured in patients without known brain metastases at the time of the PET scan (n = 293, median = 5.52) **(C)**. **(B-D)** Boxplots representation of SUVmean$_{brain}$ in the full cohort (n = 380) with brain metastases removed manually **(B)** and SUVmean$_{brain}$ measured in patients without brain metastasis only **(D)** according to the 1-year vital status. Log-rank test and Wilcoxon test *P* values are shown.

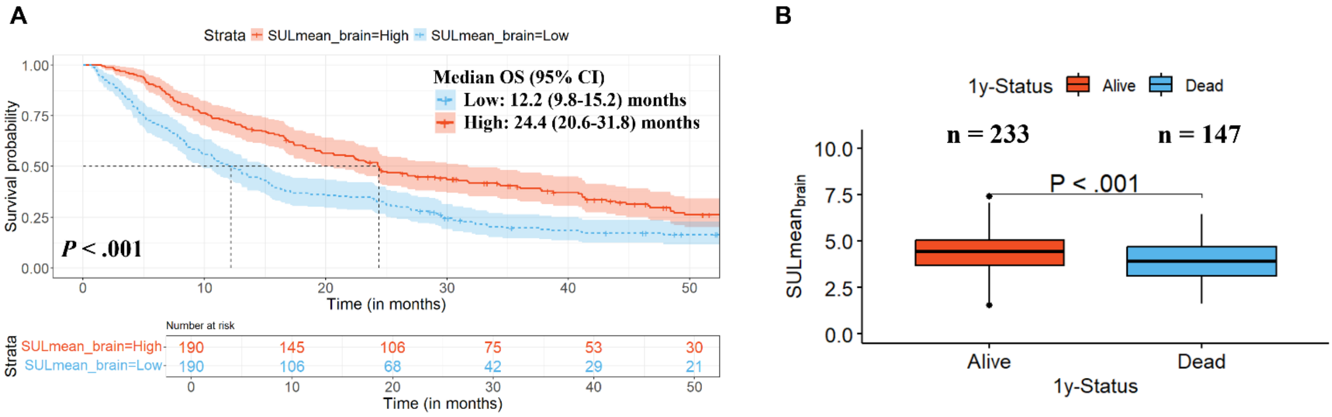


**Figure S8**: Relationship between $SULmean_{brain}$ and overall survival in metastatic NSCLC patients. **(A)** Kaplan Meier curves for NSCLC patients in the full cohort (n = 380) stratified into risk groups by the median of the $SULmean_{brain}$ (median = 4.23). Log-rank test *P* value is shown. **(B)** Boxplots representation of $SULmean_{brain}$ according to the 1-year vital status in the full cohort. Log-rank test and Wilcoxon test *P* values are shown.

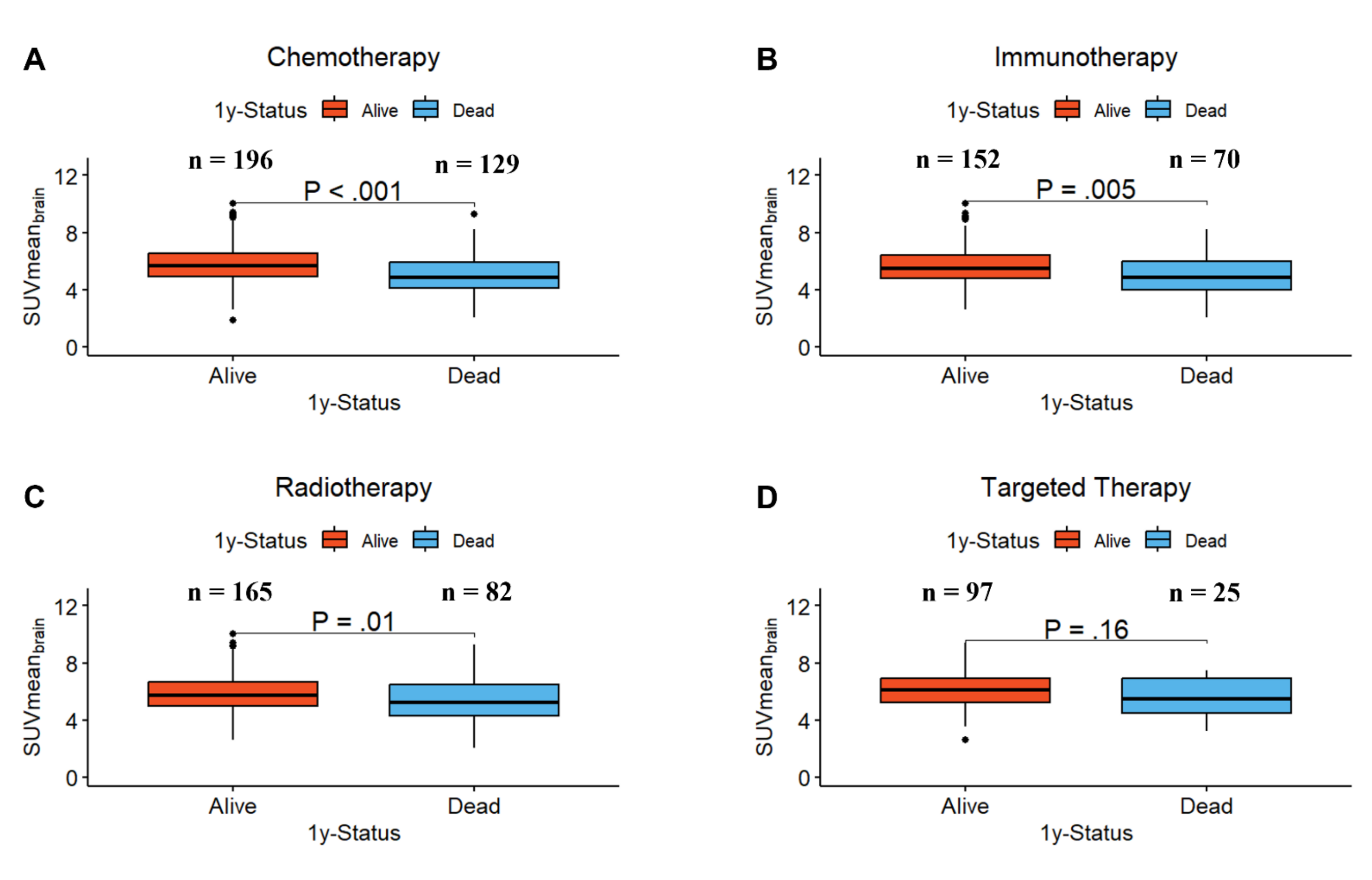


**Figure S9**: Boxplot representation of the SUVmean$_{brain}$ distribution according to the 1-year vital status for patients who received chemotherapy (n = 325/380 [86%]) **(A)**, immunotherapy (n = 222/380 [58%]) **(B)**, radiotherapy (n = 247/380 [65%]) **(C)** and targeted therapy (n = 122/380 [32%]) **(D)**. The Wilcoxon test *P* values are shown.

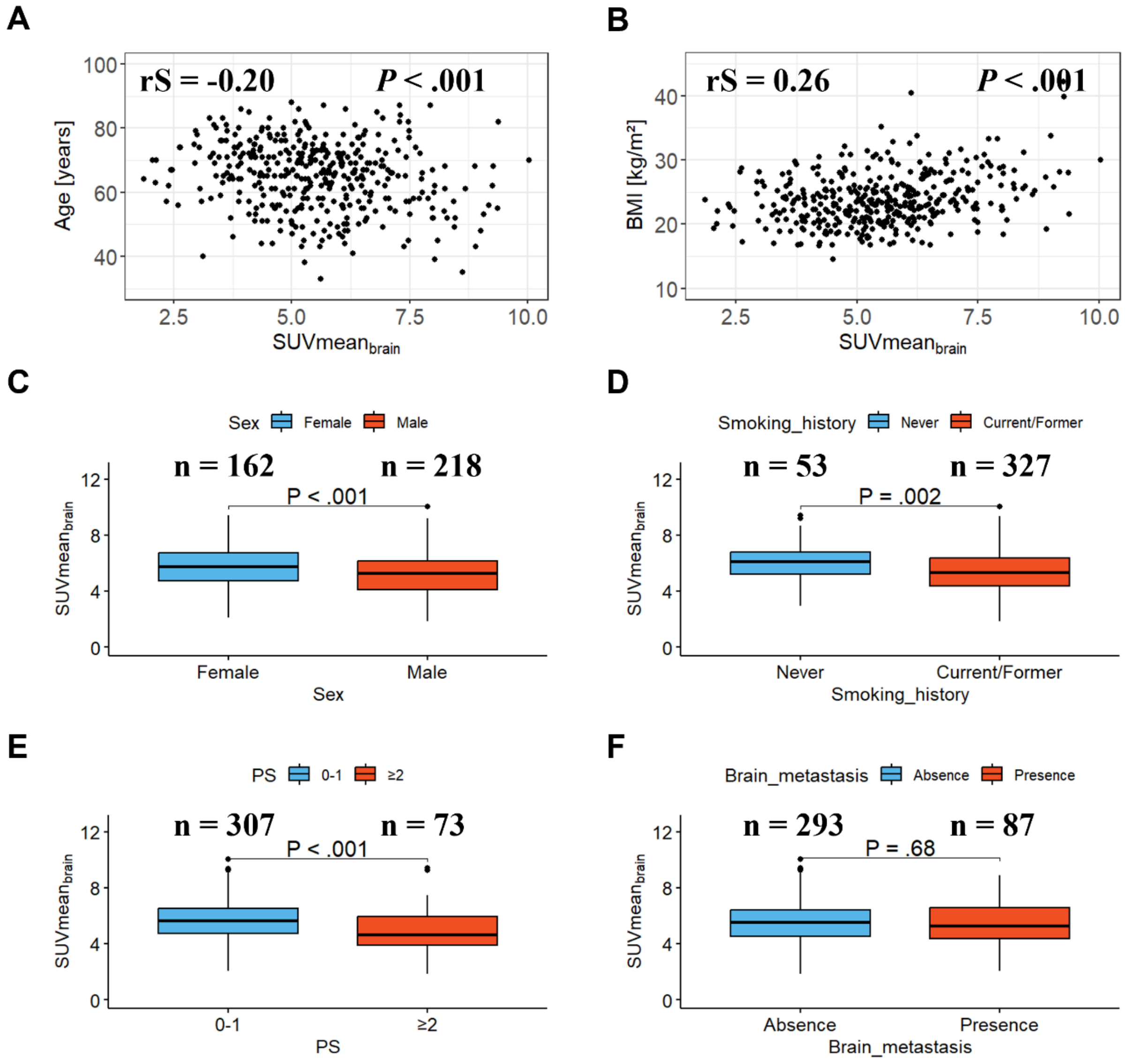


**Figure S10**: Relationship between $SUVmean_{brain}$ and clinical data using Spearman correlation for quantitative variables and boxplots representation based on Wilcoxon test for qualitative variables in the full Cohort (n = 380). **(A-B)** Correlation plot between $SUVmean_{brain}$ and Age **(A)** and Body Mass Index (BMI) **(B)**. **(C-F)** Spearman correlation coefficients (rS) and Wilcoxon test *P* values are shown. Boxplots representation between $SUVmean_{brain}$ and Sex **(C)**, Smoking history **(D)**, Performance Status (PS) **(E)** and the presence of brain metastasis **(F)**. Spearman correlation coefficients (rS) and Wilcoxon test *P* values are shown.

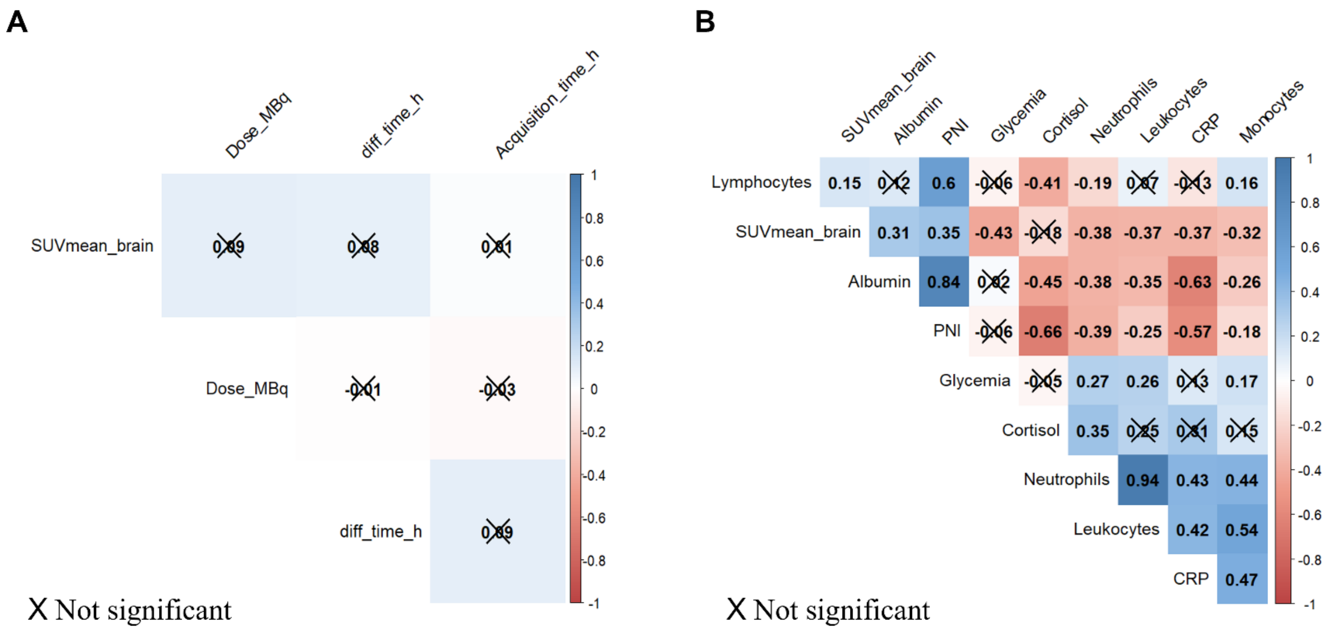


**Figure S11**: Correlation analysis between SUVmean$_{brain}$ and factors that may influence cerebral metabolic activity for the full cohort (n = 380) patients. **(A-B)** Correlogram representation showing Spearman correlation coefficients between SUVmean$_{brain}$ and technical parameters related to PET image acquisition such as the injected dose (Dose_MBq), the time from injection to acquisition (diff_time_h) and the acquisition time **(A)** and biological data **(B)**.

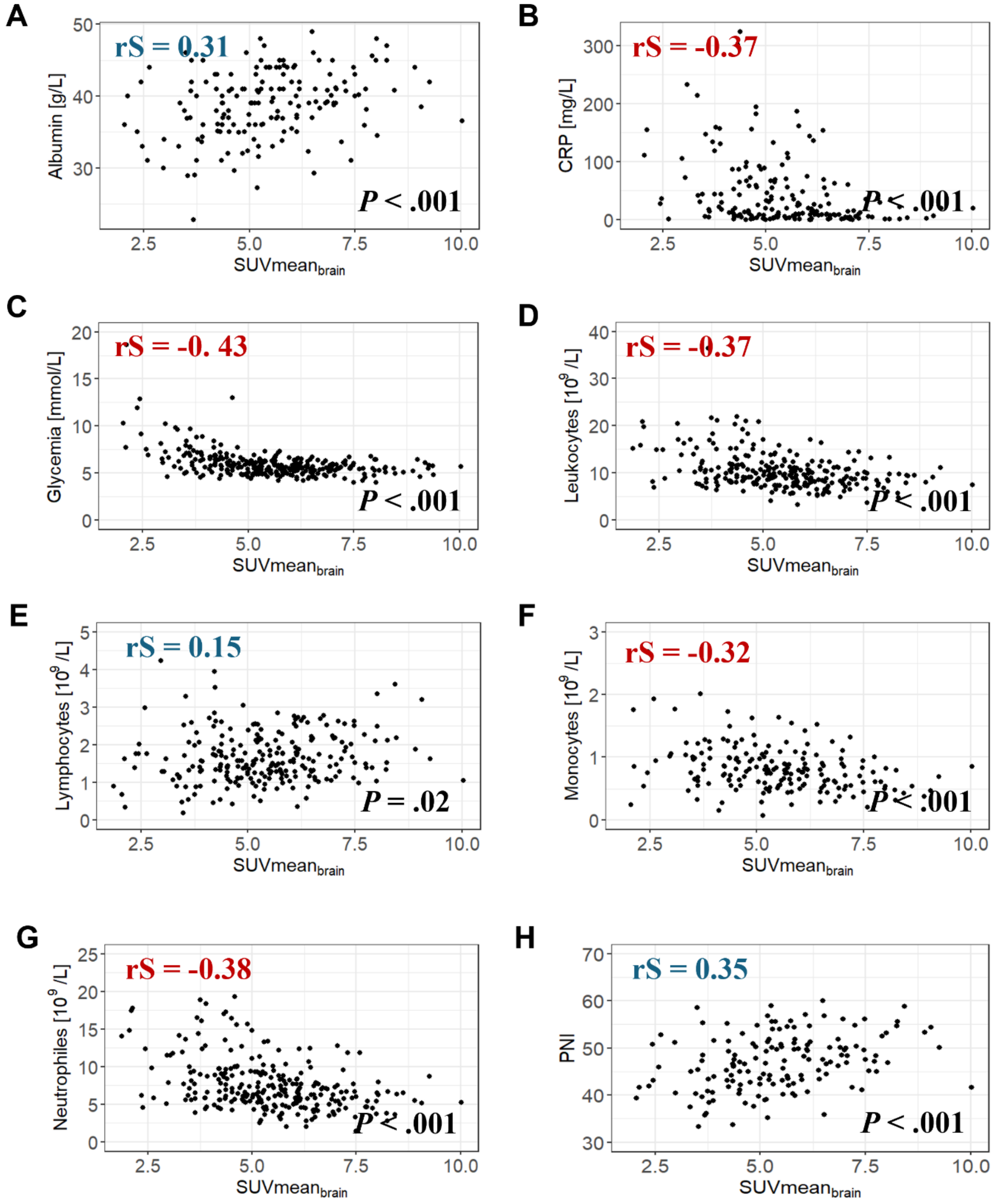


**Figure S12**: Correlation analysis between $SUVmean_{brain}$ and biological when available, among patients in the full cohort (n = 380). **(A-G)** Correlation plot between $SUVmean_{brain}$ and Albumin (n = 160) **(A)**, CRP (n = 179) **(B),** Blood glucose levels (n = 300) **(C)**, Leukocytes (n = 262) **(D)**, Lymphocytes (n = 240) **(E)**, Monocytes (n = 182) **(F)**, Neutrophils (n = 261) **(G)** and PNI (n = 149) **(H)**. Spearman correlation coefficients (rS) are shown.

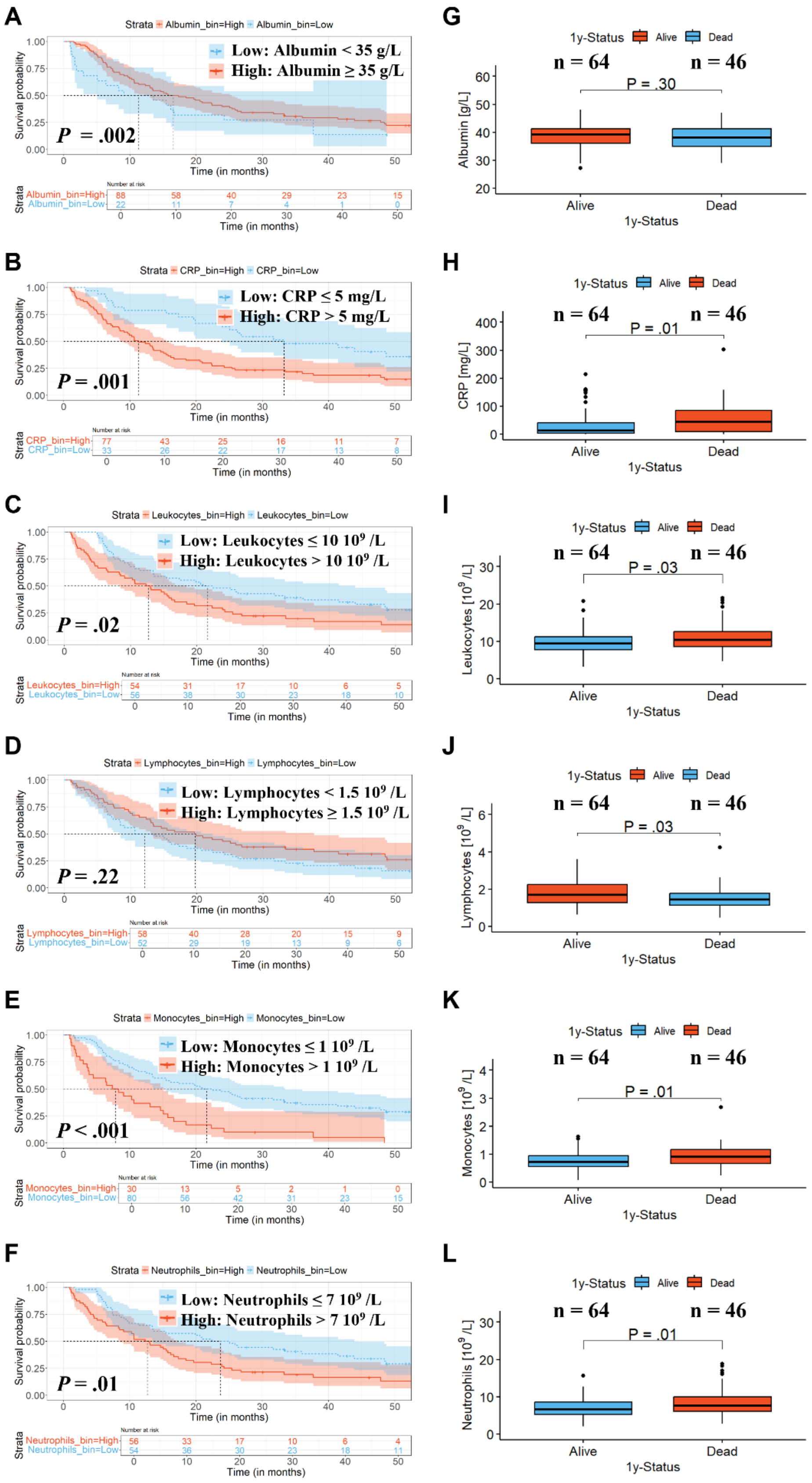


**Figure S13**: Relationship between blood biomarkers and overall survival available in a subgroup of NSCLC patients (110/380 [29%]). **(A-F)** Kaplan Meier curves for NSCLC patients stratified into risk groups based the normal biological values for the following blood biomarkers: albumin (reference = 35 g/L) **(A)**, CRP (5 mg/L) **(B)**, Leukocytes (10 $10^9$ /L) **(C)**, Lymphocytes (1.5 $10^9$ /L) **(D)**, Monocytes (1 $10^9$ /L) **(E)** and Neutrophils (7.0 $10^9$ /L) **(F)**. Log-rank test *P* value is shown. **(E-H)** Boxplots representation of Albumin **(G)**, CRP **(H)**, Leukocytes **(I)**, Lymphocytes **(G),** Monocytes **(K)** and Neutrophils **(L)** according to the 1-year vital status. Log-rank test and Wilcoxon test *P* values are shown.

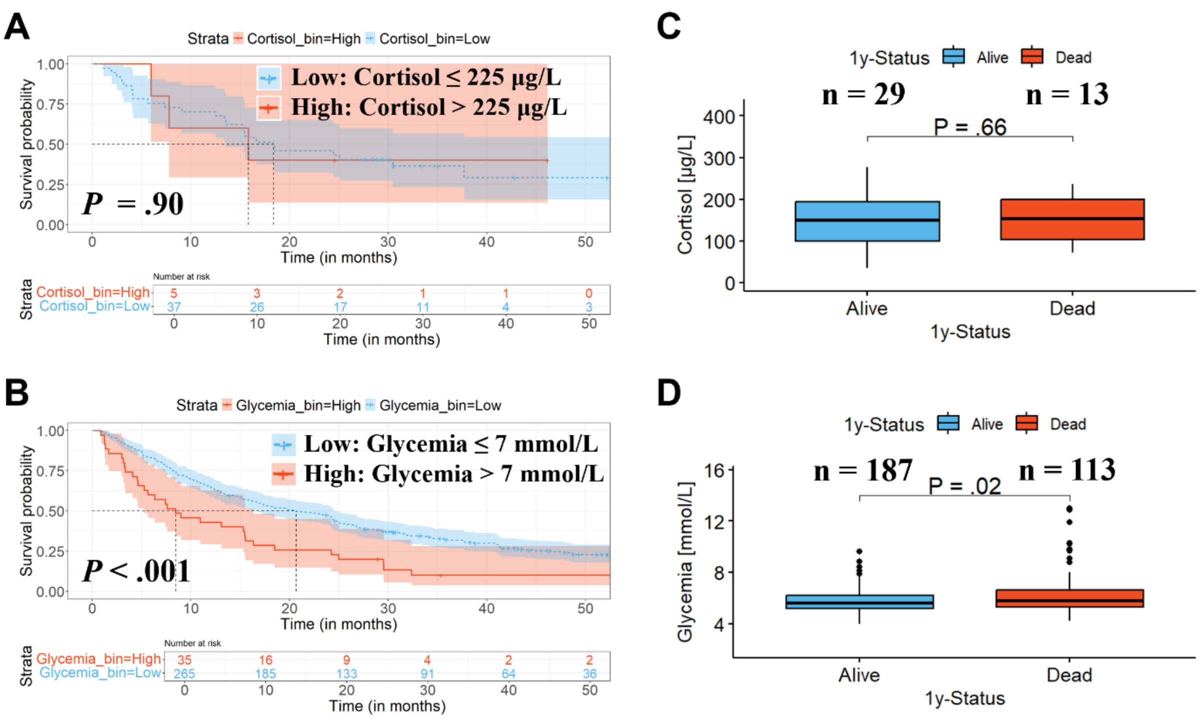


**Figure S14**: Relationship between cortisol, blood glucose levels and overall survival, when available, among patients in the full cohort (n = 380). **(A-B)** Kaplan Meier curves for NSCLC patients stratified into risk groups based the normal biological values for cortisol (n = 42) **(A)** and blood glucose levels (n = 300) **(B)**. **(C-D)** Boxplots representation of Cortisol **(C)** and Blood glucose levels **(D)** according to the 1-year vital status. Log-rank test and Wilcoxon test *P* values are shown.

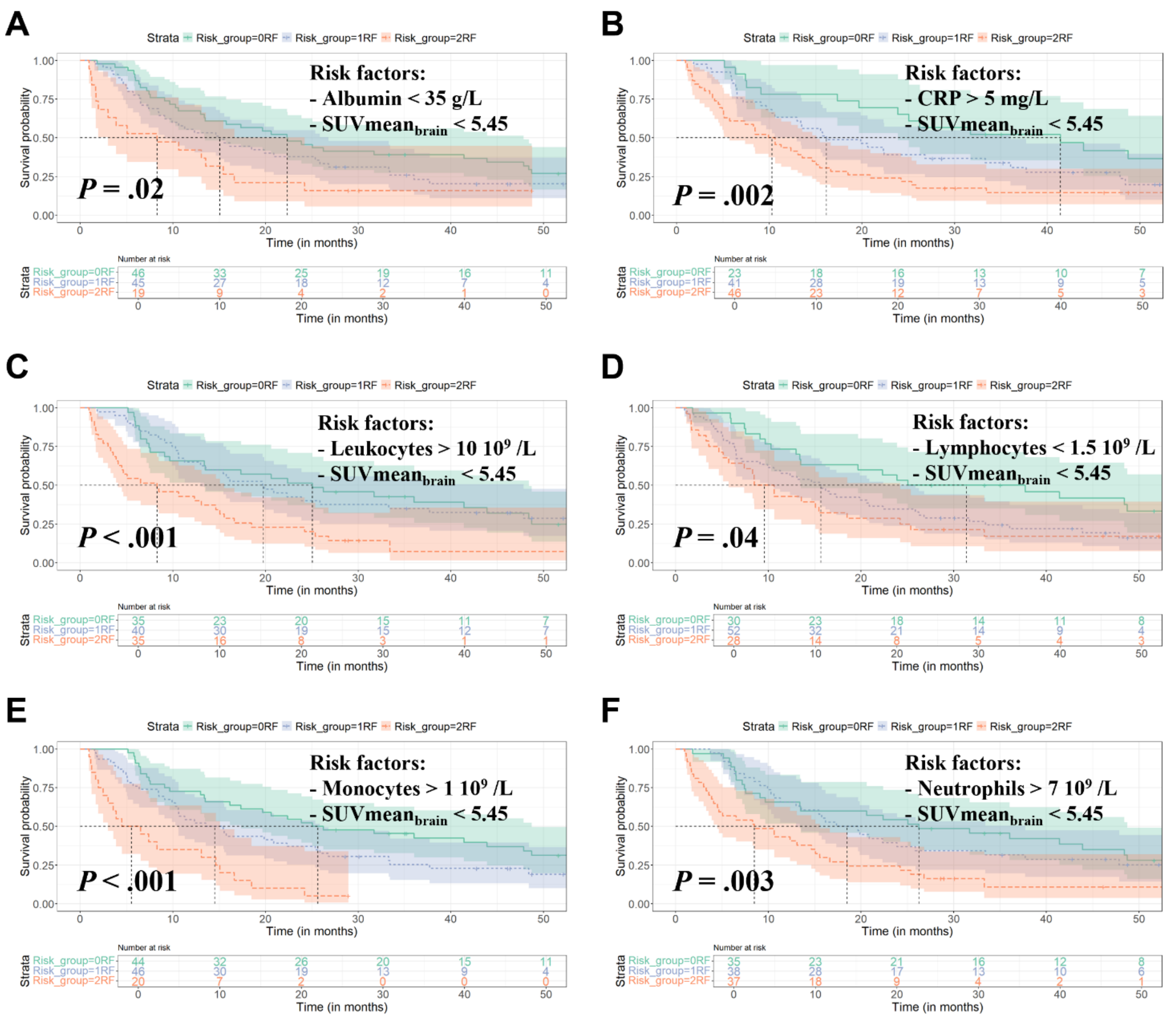


**Figure S15**: Exploration of the complementarity between SUVmean$_{brain}$, blood biomarkers and overall survival available in a subgroup of NSCLC patients (110/380 [29%]). **(A-F)** Kaplan Meier curves for NSCLC patients stratified into three risk groups (0 risk factors in green, 1 risk factor in blue and 2 risk factors in red) based on the SUVmean$_{brain}$ median value (risk factor: < 5.45) and normal biological values for the following blood biomarkers: albumin (risk factor: < 35 g/L) **(A)**, CRP (> 5 mg/L) **(B)**, Leukocytes (> 10 $10^9$ /L) **(C)**, Lymphocytes (< 1.5 $10^9$ /L) **(D)**, Monocytes (> 1.0 $10^9$ /L) **(E)** and Neutrophils (> 7.0 $10^9$ /L) **(F)**. Log-rank test *P* value is shown.

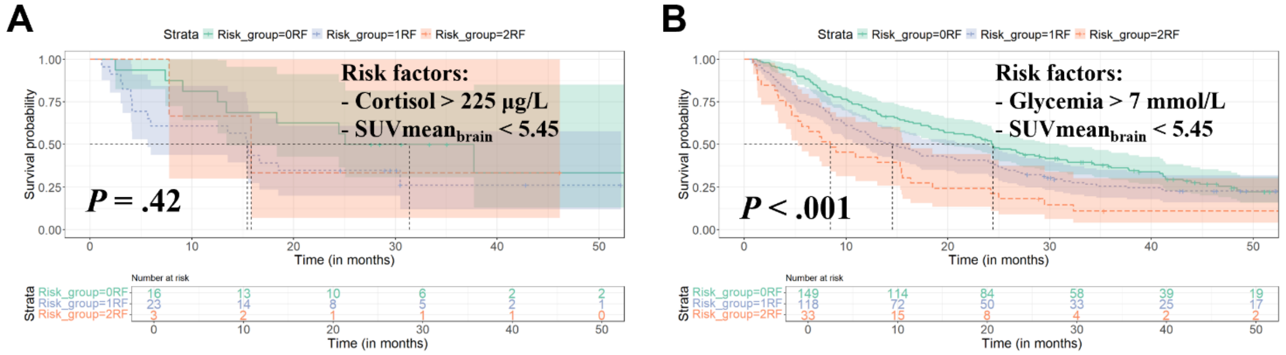


**Figure S16**: Exploration of the complementarity between SUVmean$_{brain}$, cortisol, blood glucose levels and overall survival, when available, among patients in the full cohort (n = 380). **(A-B)** Kaplan Meier curves for NSCLC patients stratified into three risk groups (0 risk factors in green, 1 risk factor in blue and 2 risk factors in red) based on the SUVmean$_{brain}$ median value (risk factor: < 5.45) and normal biological values for the following blood biomarkers: cortisol (risk factor: > 225 µg/L, n = 42) **(A)** and blood glucose levels (≥ 7 mmol/L, n = 300) **(B)**. Log-rank test *P* values are shown.

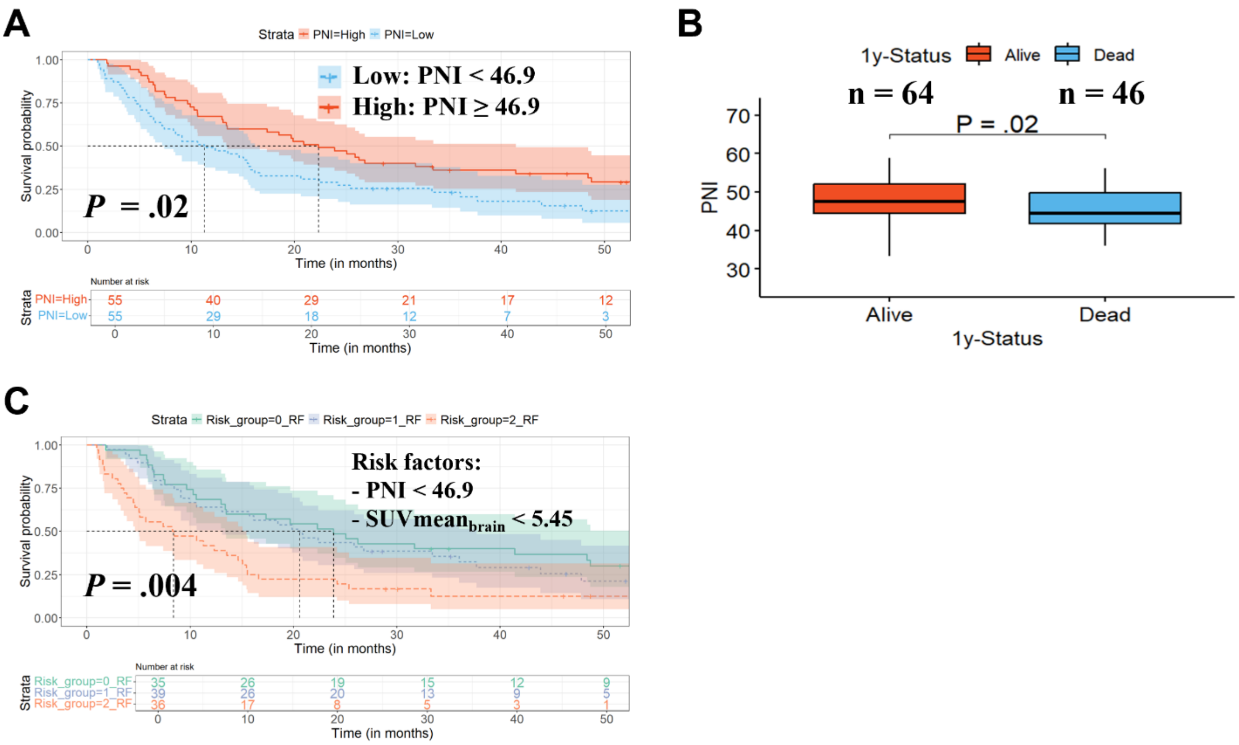


**Figure S17**: Relationship between Prognostic Nutritional Index (PNI) and overall survival, and its complementarity with SUVmean$_{brain}$ in a subgroup of NSCLC patients (110/380 [29%]). **(A)** Kaplan Meier curves for NSCLC patients stratified into risk groups based on the median PNI value. **(B)** Boxplot representation of according to the 1-year vital status. **(C)** Kaplan Meier curves for NSCLC patients stratified into three risk groups (0 risk factors in green, 1 risk factor in blue and 2 risk factors in red) based on the median values for SUVmean$_{brain}$ (< 5.45) and PNI (< 46.9). Log-rank test and Wilcoxon test *P* values are shown.

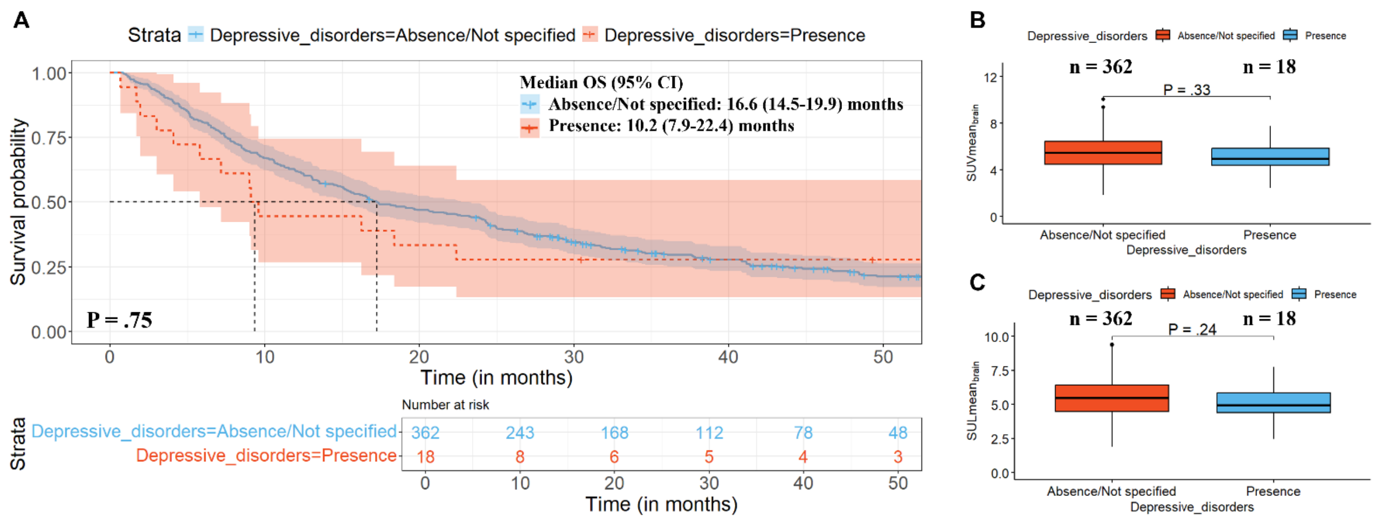


**Figure S18**: Relationship between depressive disorders, brain FDG uptake and overall survival in metastatic NSCLC patients. **(A)** Kaplan Meier curves for NSCLC patients in the full cohort (n = 380) stratified according to the presence (n = 18/380 [5%]) or absence (n = 362/380 [95%]) of depressive disorders. **(B-C)** Boxplots representation of $SUVmean_{brain}$ **(B)** and $SULmean_{brain}$ **(C)** according to the presence or absence of depressive disorders in the full cohort. Log-rank test and Wilcoxon test *P* values are shown.
